# Seasonality and spatial heterogeneity of confirmed malaria in Senegal (2009-2025): consistency with VECTRI simulations driven by ten bias-adjusted CMIP6 models and projected changes

**DOI:** 10.64898/2026.11.09.26367673

**Authors:** Ibrahima Diouf, Marie Jeanne Sambou Cissokho, Papa Fall, Amanatou Bèye Gueye, Elhadji Doucouré, Amadou Thierno Gaye

**Affiliations:** Faculty of Sciences and Techniques (FST), University of Labé, Hafia, Labé, Guinea; Laboratory of Atmospheric and Ocean Physics-Siméon Fongang (LPAO-SF), Higher Polytechnic School (ESP), Cheikh Anta Diop University (UCAD), Dakar, Senegal; African Climate Development Initiative (ACDI), University of Cape Town (UCT), Cape Town, South Africa; National Malaria Control Programme (PNLP), Rue Aimé Césaire, Fann Résidence, Dakar, Senegal

## Abstract

**Background:** Malaria transmission in Senegal is seasonal and spatially heterogeneous, but the consistency of dynamical malaria models with routine surveillance has rarely been tested with monthly subnational data.

**Methods:** We analysed monthly confirmed cases from the National Malaria Control Programme (PNLP) regional database (14 regions, 2009-2025) and reports of the 79 health districts (2016-2025). Ten bias-adjusted NEX-GDDP-CMIP6 models (1995-2014; SSP2-4.5 and SSP5-8.5 to 2100) were evaluated against CHIRPS rainfall; VECTRI was driven by MRI-ESM2-0, the model closest to the ensemble median change.

**Results:** National incidence averaged 24.5 cases per 1,000 per year without trend over 2009-2024, but increased in Kédougou, Kolda and Tambacounda, which reported 58.7 % of cases for 11.3 % of the population. The test positivity rate (TPR) decreased from 22.5 % in 2016 to 16.2 % in 2024. Based on the seasonal centre of gravity, observed cases lagged the entomological inoculation rate (EIR) simulated by VECTRI by 1.06 months (95 % CI 0.88-1.23) and the lag was positive in 13 of 14 regions, but the regional differences in timing were not reproduced. Across 45 departments, the simulated EIR was correlated with the TPR (Spearman ρ = 0.67, 95 % CI 0.47-0.79). In 2025, confirmed cases decreased by 55.7 % in the 34 districts of the long-lasting insecticidal net (LLIN) mass campaign against 9.7 % elsewhere. Under SSP5-8.5 in 2081-2100, the models projected +4.6 °C and -32 % annual rainfall, and the simulated EIR decreased by 35 % (p < 0.001).

**Conclusions:** VECTRI driven by bias-adjusted CMIP6 climate reproduces the timing of the malaria season and the gradient of transmission intensity in Senegal, but not the regional differences in timing. The TPR is a population-free indicator for model evaluation. Seasonal malaria chemoprevention calendars should account for the regional timing documented here; the projected changes in timing are small.

## Introduction

Malaria remains a major public health problem in sub-Saharan Africa, which bears most of the global burden of cases and deaths [1]. In Senegal, transmission is linked to the West African monsoon: it is concentrated in the rainy season and follows a south-east to north gradient [2–4]. This seasonality underlies seasonal malaria chemoprevention (SMC), recommended by the World Health Organization for areas of highly seasonal transmission in the Sahel [5] and shown to be effective in children under ten years of age in Senegal [6].

Weather-driven dynamical models, such as the Liverpool Malaria Model (LMM) [7–9] and VECTRI [10,11], simulate vector and parasite development as a function of rainfall and temperature and produce indicators such as the entomological inoculation rate (EIR). They have been used over Senegal and West Africa to reconstruct past transmission [2,3,12], to relate malaria variability to climate and ocean conditions [13,14], to assess seasonal predictability [15–17] and to project the impact of climate change [4,18–21]. Temperature affects transmission non-linearly, with an optimum near 25 °C [22], so that rainfall and temperature jointly control the length and intensity of the season. In Senegal, simulations place the peak of transmission in September-October, one to two months after the rainfall peak, reflecting the time required for mosquito breeding and the sporogonic cycle. [2–4,13,19].

Two limitations recur in these studies. First, their evaluation against observed malaria has relied on short periods, sentinel sites or annual totals [2,20], whereas the National Malaria Control Programme (PNLP) now reports monthly data for all health districts, including the number of suspected and tested patients, which allows the test positivity rate (TPR) to be computed [23]. Second, climate change projections were often driven by one model or by a multi-model mean forcing, which does not represent the spread of the projections and, because the response of malaria models is non-linear, does not correspond to the mean of single-model simulations. The NASA Earth Exchange Global Daily Downscaled Projections (NEX-GDDP-CMIP6) provide daily rainfall and temperature from many models of the Coupled Model Intercomparison Project Phase 6 (CMIP6) [24], bias-adjusted and downscaled to 0.25° [25], which allows the spread of the climate projections to be described and a representative model to be selected to drive a malaria model.

This study has four objectives: (i) to describe the seasonality, spatial distribution, trends and severity of confirmed malaria in Senegal from the PNLP regional database (2009-2025) and harmonised district reports (2016-2025); (ii) to evaluate the bias-adjusted CMIP6 rainfall against observations and the timing and spatial pattern of transmission simulated by VECTRI against surveillance data, at the national, regional and departmental scales; (iii) to describe the 2025 surveillance data in relation to the LLIN mass distribution campaign; and (iv) to project changes in climatic suitability and simulated transmission under two emission scenarios.

## Materials and methods

### Study area

Senegal (about 197,000 km^2^) is divided into 14 regions and 45 departments (GADM version 4.1 boundaries, https://gadm.org) and into 79 health districts. Annual rainfall decreases from more than 1,100 mm in the south (Ziguinchor, Kédougou) to less than 300 mm in the north (Saint-Louis), with a single rainy season from June to October peaking in August. The analysis is conducted on a regular 0.25° grid (331 cells overlapping Senegal), with the share of each cell inside each region and department used as aggregation weight together with the cell area.

### Malaria surveillance data

Two PNLP sources were used (Fig 1). The **regional database** provides monthly confirmed malaria cases (rapid diagnostic test or microscopy) and malaria deaths for the 14 regions from 2008 to 2025, with the regional population of the National Agency of Statistics and Demography (ANSD). The year 2008 was excluded because of the change of regional boundaries, and 2025 is provisional; the analysis period is 2009-2024 (16 years), and 2025 is reported separately.

**Fig 1.**
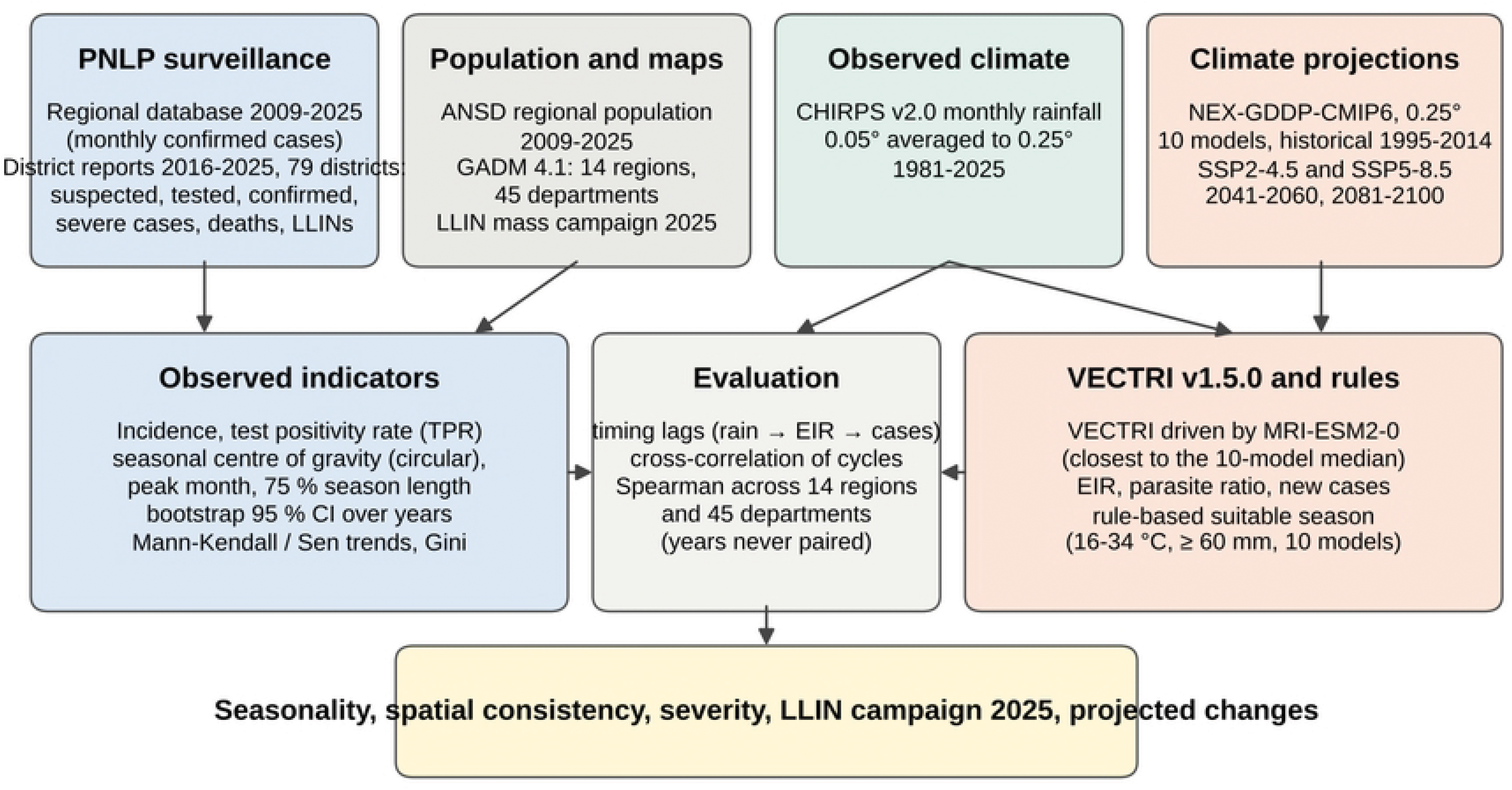
Data sources and analysis workflow. PNLP: National Malaria Control Programme; ANSD: National Agency of Statistics and Demography; LLIN: long-lasting insecticidal net; EIR: entomological inoculation rate; TPR: test positivity rate.

The **district reports** were harmonised from the annual Excel reports of 2016-2020 and the District Health Information Software 2 (DHIS2) exports of 2021-2025. They provide, by district and month, the number of suspected, tested and confirmed outpatient cases for children under five years, patients aged five years and over (excluding pregnant women) and pregnant women, the number of hospitalised malaria cases (hospitalised severe malaria in 2016-2019, hospitalised confirmed malaria from 2020) and malaria deaths, and, for 2021-2025, the number of LLINs distributed through routine channels. In the 2016-2019 reports, 36 to 37 hospitals were reported as separate units; they were added to the district in which they are located, as in the 2020 report and the DHIS2 exports, which integrate hospitals in their district. Districts created after 2016 (Sangalkam, Yeumbeul and Diakhao) have no separate report before their creation; analyses over 2016-2025 were therefore made by department (GADM level 2, each district being assigned to the department where it lies) or by region. All reporting-unit names were matched. The district reports also provide the number of LLINs distributed during the 2025 mass distribution campaign (CDM) in 34 districts.

In the DHIS2 exports, 235 of the 3,792 district-months of 2021-2024 (6.2 %) had no value for confirmed cases although the number of tested patients was reported (mean 720 tests); 94.5 % of them fell between January and June, mostly in low-transmission districts of Louga, Fatick and Matam. Because DHIS2 does not store zero values, they were counted as zero. Monthly report completeness, available for 2017-2020, ranged from 95.7 % to 99.6 %.

Annual national totals of the two sources were identical for 2016-2020 and differed by -0.2 % (2021), +0.0 % (2023) and -0.1 % (2024); in 2022, the regional database was 21.8 % lower (358,033 against 458,040 confirmed cases), with lower values in all regions (S1 Fig). The regional database is used for the long period and the district reports for testing, positivity, age, severity and departmental analyses; the sensitivity of the main result to 2022 and to the source was assessed.

### Observed rainfall

Monthly rainfall from the Climate Hazards Group InfraRed Precipitation with Station data (CHIRPS v2.0, 0.05°) [26] for 1981-2025 was averaged to the 0.25° grid (mean of the 25 CHIRPS cells of each 0.25° cell) and aggregated to regions and departments.

### CMIP6 climate projections

Daily precipitation and near-surface air temperature of ten CMIP6 models from NEX-GDDP-CMIP6 [25] were used: ACCESS-CM2, CNRM-CM6-1, EC-Earth3, GFDL-ESM4, INM-CM5-0, IPSL-CM6A-LR, MIROC6, MPI-ESM1-2-HR, MRI-ESM2-0 and NorESM2-MM, one per modelling centre, for the historical experiment (1995-2014) and the SSP2-4.5 and SSP5-8.5 scenarios [27] (2041-2060 and 2081-2100). CanESM5, a model with very high climate sensitivity [28], and the second members of the ACCESS and CNRM families were not included. NEX-GDDP-CMIP6 is bias-adjusted against the Global Meteorological Forcing Dataset [29] and has no data over the ocean; the 53 Atlantic cells of the box were excluded, and all 331 cells overlapping Senegal had complete data. Missing days of non-standard calendars (360-day and 365-day) were filled by linear interpolation for temperature and with zero for rainfall before computing monthly values. The historical rainfall of each model was compared with CHIRPS over 1995-2014 (annual bias, correlation of the mean seasonal cycle and difference of rainfall centre of gravity).

To drive VECTRI, one model was selected objectively: for each scenario and period, the changes in annual rainfall (%), July-September rainfall (%) and mean temperature (°C) over Senegal were scaled by the spread of the models (median absolute deviation), and the model whose changes were on average closest to the ensemble median was retained. MRI-ESM2-0 [30] ranked first (distance 0.86), followed by MIROC6 (0.99) and GFDL-ESM4 (1.40); EC-Earth3 (5.28) and INM-CM5-0 (4.49), the two models projecting wetter conditions, ranked last.

### VECTRI simulations

VECTRI version 1.5.0 [10,11] is a grid-based dynamical malaria model that represents the effects of rainfall on temporary breeding sites, of temperature on larval development, adult survival and the sporogonic cycle, of population density on biting rates, and a simple representation of immunity; default parameters were used, without calibration and without interventions. VECTRI was run on the 0.25° grid with the daily MRI-ESM2-0 forcing for 1995-2014 and the four future periods, each preceded by a five-year spin-up (the first years of the period replayed) that was removed from the results, and with the population density of AfriPop [31] interpolated bilinearly from 1° to 0.25°. Monthly EIR (infective bites per person per month), new cases per person, detectable parasite ratio, rainfall and temperature were computed from daily outputs and aggregated to the national, regional and departmental units with area weights. The transmission season length was defined as the longest run of consecutive months with EIR ≥ 1 infective bite per person per month in each cell.

### Rule-based climatic suitability

As a model-independent complement driven by all ten models, a month was considered climatically suitable when its mean temperature was between 16 and 34 °C [22] and its rainfall reached 60 mm, provided that at least one month of the twelve-month window reached 80 mm [32,33]; the suitable season is the longest run of suitable months in a twelve-month window starting at the driest month of each cell, and suitability is stable where the mean season exceeds three months [33,34]. Changes were computed for each model and summarised by the ensemble median, the range and the number of models with a significant change (Mann-Whitney test on annual values, p < 0.05); model agreement on the sign of the change was mapped where at least 8 of the 10 models agreed.

### Indicators and statistical analysis

Annual incidence was expressed as confirmed cases per 1,000 inhabitants (‰) and the TPR as confirmed cases per tested patient. Because more than 96 % of suspected cases were tested each year (see Results), the TPR measures the share of febrile patients with a malaria infection and is independent of population estimates; it is less sensitive than incidence to care seeking, although it depends on the background of non-malarial fevers [23]. Seasonal timing was summarised with circular statistics [35]: months were represented as angles (30° per month) weighted by their number of cases, giving the circular mean month, or centre of gravity (9 = mid-September, 10 = mid-October), and the concentration R (0 = cases evenly distributed over the year, 1 = all cases in one month). The centre of gravity was computed for each year and averaged as a circular mean, with a 95 % CI from 2,000 bootstrap resamples of years [36]. The season length was described by the shortest run of consecutive months holding 75 % of the annual total and by the contiguous months reaching 20 % of the peak of the mean cycle. Monotonic trends were tested with the Mann-Kendall test [37,38] and quantified with the Sen slope [39]. The spatial concentration of cases relative to population was summarised by the Gini coefficient across regions. Severity was described by malaria deaths per 1,000 confirmed cases with exact Poisson CIs [40]; this ratio is not a case fatality rate because deaths are not linked to individual outpatient cases.

The comparison between observations and simulations relies on climatological seasonality only: CMIP6 historical simulations are free-running and do not reproduce the observed chronology of years, so simulated and observed years were never paired. The lag of observed cases behind simulated transmission was estimated as the circular difference between the mean centres of gravity of observed cases (2009-2024) and of the simulated EIR (1995-2014), with a CI obtained by resampling observed and simulated years independently. As a shape-based estimate, the Pearson correlation between the normalised mean cycles (monthly share of the annual total) was computed for lags of 0 to 4 months. The same statistics were computed between observed cases and CHIRPS rainfall (2009-2024) and between the simulated EIR and the simulated rainfall, which separates the delay between rainfall and transmission, reproduced by the model, from the delay between transmission and reported cases. Spatial correspondence was assessed with Spearman correlations across the 14 regions and 45 departments, with bootstrap CIs over units. Regional and departmental analyses are exploratory; where several tests are reported, a Bonferroni threshold is indicated.

Projected changes in the simulated EIR were tested with the Mann-Whitney test on annual values (20 years per period), and changes in the EIR centre of gravity with bootstrap CIs. The 2025 data of the districts covered by the LLIN mass campaign were compared with those of the other districts, as the ratio of 2025 to the mean of 2021-2024 for confirmed cases, tested patients and TPR (Mann-Whitney test across districts); this comparison is descriptive, because campaign districts were selected for their high burden and the 2025 data are provisional.

### Software, reproducibility and ethics

The chain (download, preparation, VECTRI runs, suitability analysis and dashboard) runs in Python (NumPy, SciPy, netCDF4, pandas, Matplotlib) and is published with an open interactive dashboard that presents all indicators by region and department [41]. This study used aggregated surveillance data (monthly counts by health district and region) provided by the PNLP, without individual information.

## Results

### Burden, trends and surveillance indicators

Over 2009-2024, national confirmed cases ranged from 174,890 (2009) to 536,850 (2021), and national incidence from 14.5 ‰ (2009) to 34.3 ‰ (2015), with a mean of 24.5 ‰ and no monotonic trend (Kendall τ = 0.08, p = 0.69; Sen slope +0.12 ‰ per year, 95 % CI -0.51 to 0.96) (Fig 2b; Fig 3a, 3b). The burden was strongly concentrated (Gini = 0.60; S2 Fig): Kédougou (mean 293.2 ‰), Kolda (114.8 ‰) and Tambacounda (91.9 ‰) reported 58.7 % of the cases for 11.3 % of the population, and incidence ranged from 1.7 ‰ in Saint-Louis to 293.2 ‰ in Kédougou. Incidence increased in these three regions (Kédougou τ = 0.62, p < 0.001, Sen slope +30.3 ‰ per year, 95 % CI 16.4 to 39.4; Kolda τ = 0.45, p = 0.015, +9.6 ‰ per year; Tambacounda τ = 0.43, p = 0.020, +4.9 ‰ per year) and decreased in Kaffrine, Dakar, Thiès, Fatick and Saint-Louis (τ between -0.57 and -0.48, p between 0.002 and 0.009) (S3 Fig). With a Bonferroni threshold for 14 tests (p < 0.0036), the increase in Kédougou and the decrease in Kaffrine remain significant.

**Fig 2.**
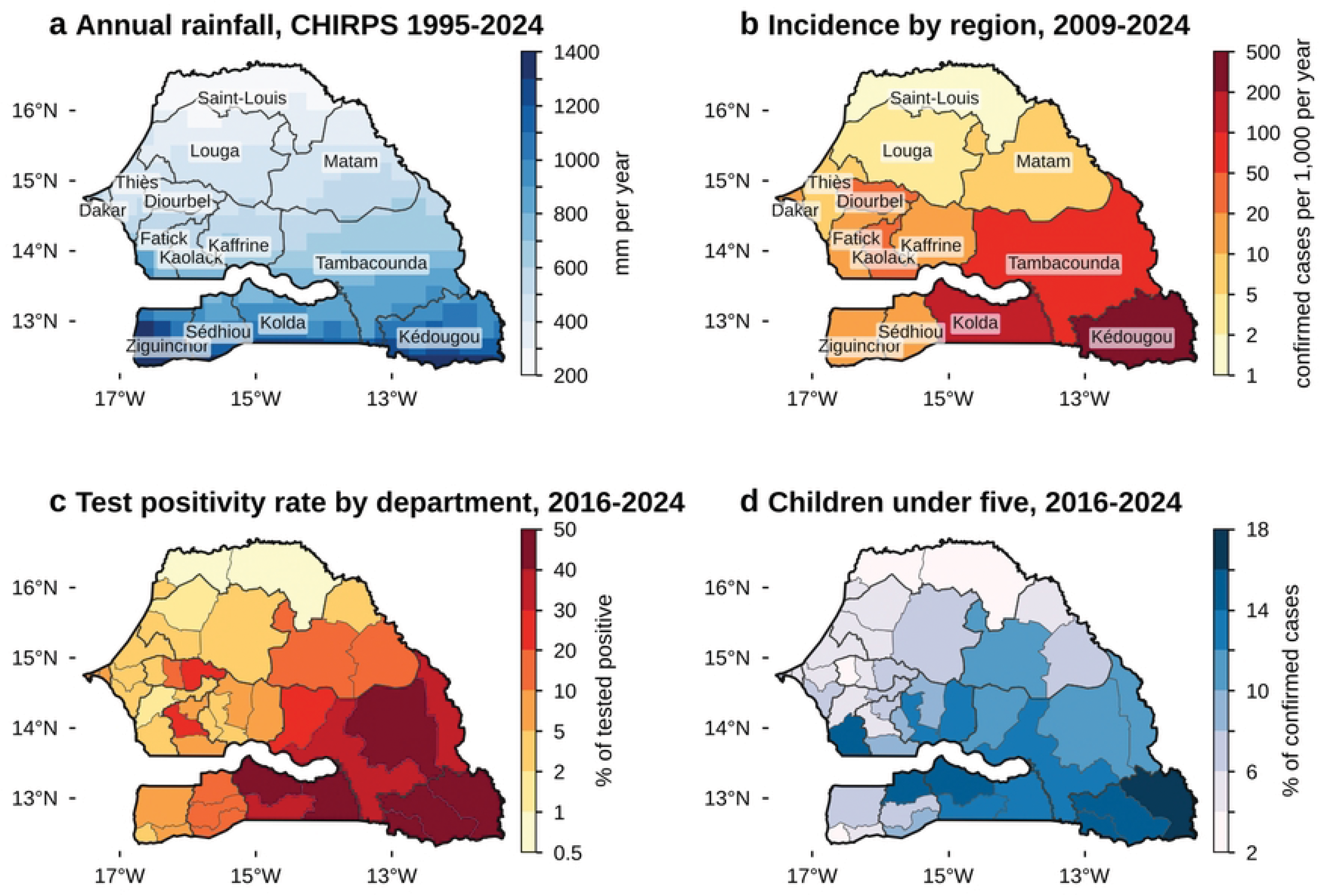
Rainfall and malaria burden in Senegal. (a) Mean annual rainfall, CHIRPS 1995-2024, 0.25° grid. (b) Mean annual incidence of confirmed malaria by region, PNLP regional database, 2009-2024 (log classes). (c) Test positivity rate by department, district reports 2016-2024. (d) Share of children under five among confirmed cases by department, 2016-2024. Region boundaries in black, department boundaries in grey (GADM 4.1).

**Fig 3.**
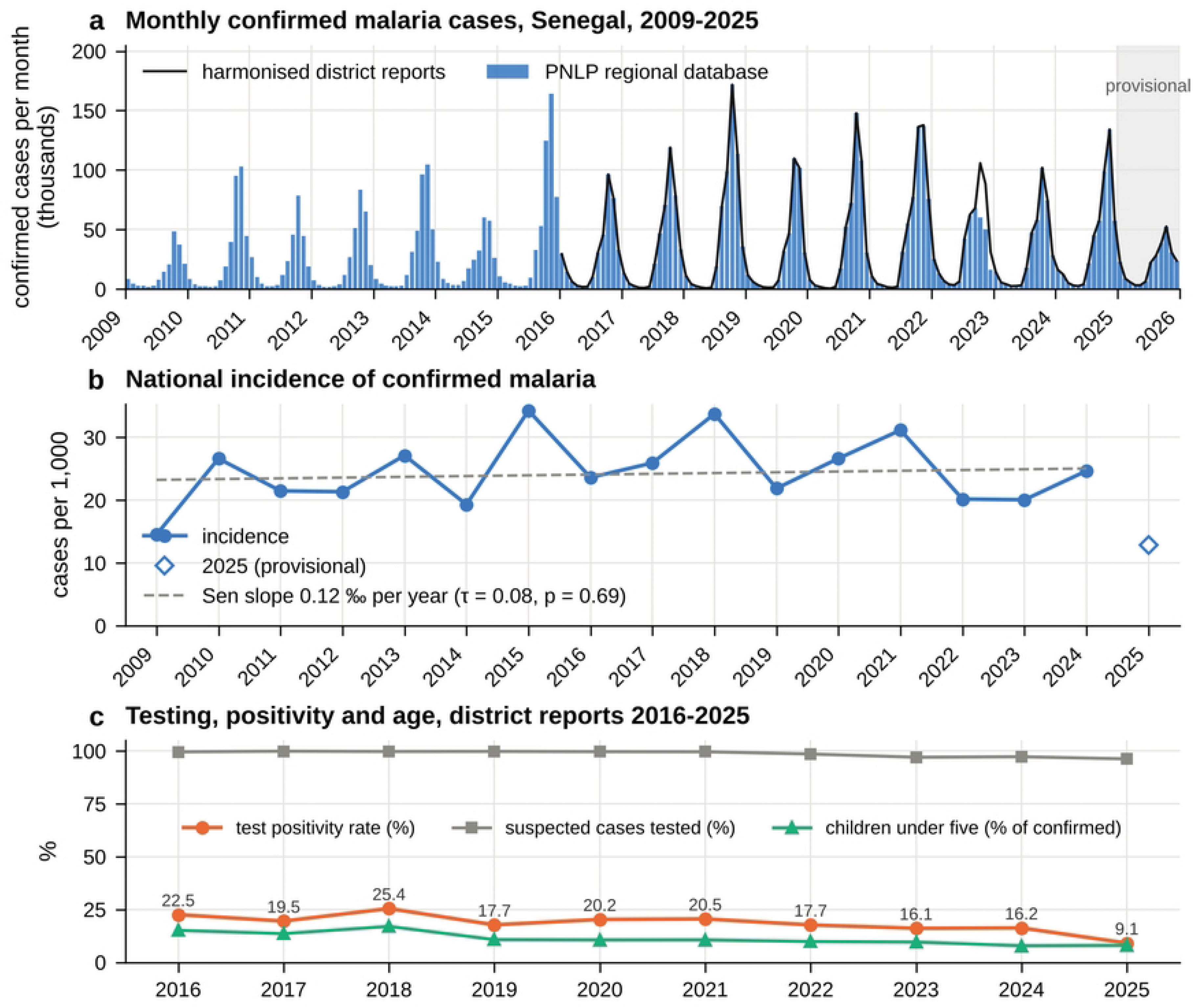
National malaria series. (a) Monthly confirmed cases from the PNLP regional database (bars, 2009-2025) and from the harmonised district reports (black line, 2016-2025); shading: provisional 2025 data. The two sources differ mainly in 2022. (b) Annual incidence with Sen slope over 2009-2024 (2025 provisional, not included). (c) Share of suspected cases tested, test positivity rate and share of children under five among confirmed cases, district reports 2016-2025.

In the district reports, 96.2 % to 99.9 % of suspected cases were tested each year (Table 1; Fig 3c). The TPR decreased from 22.5 % in 2016 and 25.4 % in 2018 to 16.1-16.2 % in 2023-2024 (τ = -0.56, p = 0.045; Sen slope -0.80 percentage point per year, 95 % CI -1.61 to -0.01), while the number of tests per inhabitant did not change significantly (τ = 0.33, p = 0.26). The share of children under five among confirmed cases decreased from 15.1 % (2016) to 7.8 % (2024). Across the 45 departments, the TPR over 2016-2024 ranged from 0.8 % (Dagana) to 58.3 % (Saraya) (Fig 2c), and it was closely related to incidence across regions (Spearman ρ = 0.90, p < 0.001). The share of children under five increased with the TPR across departments (ρ = 0.70, 95 % CI 0.46 to 0.85), from 2-5 % in low-positivity departments of the north and west to 14-17 % in the south-east (Fig 2d).

**Table 1.** National surveillance indicators from the harmonised district reports, Senegal, 2016-2025.

| Year | Suspected | Tested (%) | Confirmed | TPR (%) | Under five (%) | Incidence (‰) | Hospitalised | Deaths | Deaths per 1,000 confirmed |
|---|---|---|---|---|---|---|---|---|---|
| 2016 | 1,559,054 | 99.6 | 349,540 | 22.5 | 15.1 | 23.6 | 9,918 | 325 | 0.93 |
| 2017 | 2,035,693 | 99.9 | 395,706 | 19.5 | 13.5 | 25.9 | 10,463 | 284 | 0.72 |
| 2018 | 2,096,124 | 99.7 | 530,944 | 25.4 | 17.0 | 33.8 | 13,350 | 555 | 1.05 |
| 2019 | 2,010,398 | 99.8 | 354,708 | 17.7 | 10.7 | 21.9 | 9,352 | 260 | 0.73 |
| 2020 | 2,206,842 | 99.7 | 445,313 | 20.2 | 10.6 | 26.7 | 9,179 | 373 | 0.84 |
| 2021 | 2,635,327 | 99.6 | 538,088 | 20.5 | 10.6 | 31.2 | 15,700 | 399 | 0.74 |
| 2022 | 2,632,620 | 98.5 | 458,040 | 17.7 | 9.8 | 25.8 | 15,017 | 340 | 0.74 |
| 2023 | 2,323,432 | 97.0 | 361,877 | 16.1 | 9.6 | 20.1 | 12,431 | 297 | 0.82 |
| 2024 | 2,909,574 | 97.2 | 459,311 | 16.2 | 7.8 | 24.7 | 15,421 | 274 | 0.60 |
| 2025 * | 2,830,010 | 96.2 | 247,179 | 9.1 | 8.0 | 13.0 | 11,876 | 155 | 0.63 |

*TPR: test positivity rate (confirmed / tested). Hospitalised: hospitalised severe malaria (2016-2019, hospital reports) and hospitalised confirmed malaria (2020-2025). Deaths: malaria deaths reported by hospitals (2016-2020) and in DHIS2 (2021-2025); the two series are not strictly comparable. Incidence uses the ANSD population. \* 2025 provisional.*

### Seasonality of testing, confirmed cases and severe malaria

Nationally, the peak of confirmed cases occurred in October in 10 of the 16 years of 2009-2024, in November in five years and in September in one year. The seasonal centre of gravity was 10.14 (95 % CI 10.00-10.27), between 9.40 (2022) and 10.52 (2015), with a concentration R of 0.68; 75 % of annual cases occurred within four consecutive months (median over years), and 75.3 % (95 % CI 71.9-78.6 %) occurred between August and November (S4 Fig). The seasonality of confirmed cases was not driven by testing (Fig 4a): over 2016-2024, the number of tested patients varied only 4.8-fold between the lowest and the highest month of the mean cycle (R = 0.36) against 67-fold for confirmed cases (R = 0.69), and the TPR rose from 2.0 % in May to 33.6 % in November (Fig 4b). Hospitalised cases followed outpatient confirmed cases by 0.21 month (95 % CI 0.04 to 0.38; centre of gravity 10.28), and malaria deaths had the same timing as confirmed cases (10.19; difference 0.12 month, -0.10 to 0.33).

**Fig 4.**
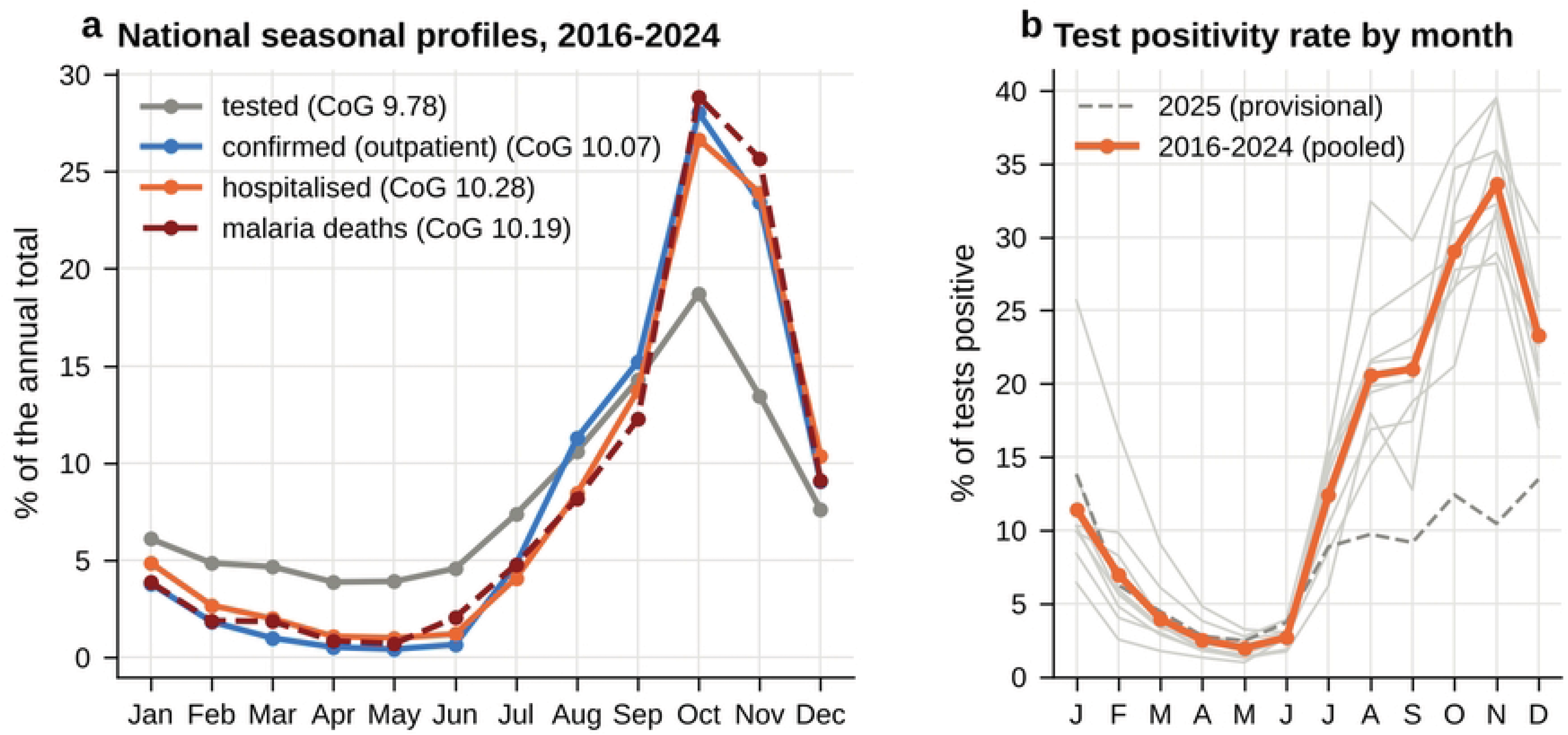
Seasonality of testing, confirmed and severe malaria, Senegal, 2016-2024. (a) Mean monthly share of the annual total of tested patients, confirmed outpatient cases, hospitalised cases and malaria deaths, with their centres of gravity (CoG, circular mean month). (b) Monthly test positivity rate: each year 2016-2024 (grey), pooled 2016-2024 (orange) and 2025 (provisional, dashed).

The timing differed between regions (Fig 5a; Table 2). The season was earliest in Kédougou (centre of gravity 9.17, 95 % CI 9.01-9.35; peak in August in 11 of 16 years), Matam (9.62) and Tambacounda (9.77), and latest in Diourbel (10.88), Sédhiou (10.85), Ziguinchor (10.74), Kaolack (10.63) and Fatick (10.62), where the peak occurred in November in up to 15 of 16 years (Ziguinchor). Across departments, the centre of gravity ranged from 8.88 to 11.09. The timing was mostly stable between 2009-2014 and 2015-2024 (national 10.25 and 10.08), but it became later in Kédougou (8.83 to 9.37) and earlier in Fatick (11.01 to 10.40) and Kaffrine (10.81 to 10.22).

**Fig 5.**
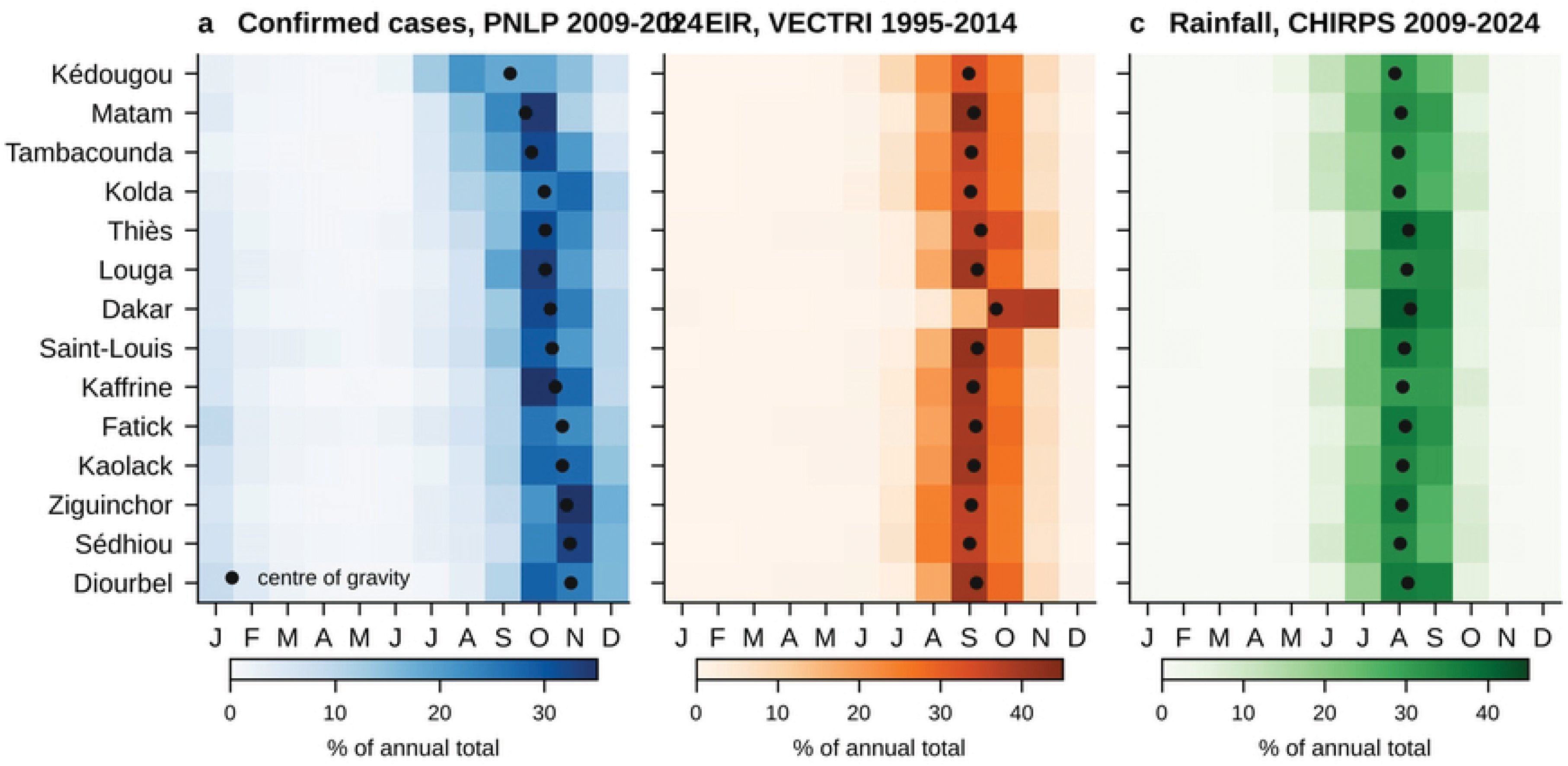
Regional seasonal cycles. Monthly share of the annual total of (a) confirmed cases (PNLP regional database, 2009-2024), (b) EIR simulated by VECTRI driven by MRI-ESM2-0 (1995-2014) and (c) CHIRPS rainfall (2009-2024). Regions are ordered by the centre of gravity of confirmed cases (black dots).

**Table 2.** Regional indicators of confirmed malaria and comparison with VECTRI.

| Region | Incidence 2009-2024 (‰), mean (range) | Trend, Sen slope ‰/year (τ; p) | TPR 2016-2024 (%) | Under five (%) | Centre of gravity, cases (95 % CI) | Centre of gravity, EIR | Lag, months (95 % CI) | Deaths per 1,000 confirmed, 2021-2024 (95 % CI) |
|---|---|---|---|---|---|---|---|---|
| Kédougou | 293.2 (58.2-534.3) | +30.30 (0.62; < 0.001) | 54.1 | 15.4 | 9.17 (9.01-9.35) | 8.99 | 0.18 (-0.02 to 0.38) | 0.31 (0.25-0.37) |
| Kolda | 114.8 (21.4-245.0) | +9.56 (0.45; 0.015) | 43.3 | 13.7 | 10.13 (9.94-10.30) | 9.03 | 1.11 (0.89 to 1.31) | 0.40 (0.34-0.46) |
| Tambacounda | 91.9 (40.7-191.3) | +4.91 (0.43; 0.020) | 37.1 | 12.2 | 9.77 (9.58-9.91) | 9.06 | 0.71 (0.48 to 0.90) | 0.50 (0.43-0.58) |
| Diourbel | 24.6 (8.6-46.0) | -0.23 (-0.08; 0.690) | 19.9 | 7.1 | 10.88 (10.66-11.09) | 9.19 | 1.69 (1.43 to 1.94) | 0.77 (0.66-0.89) |
| Kaolack | 22.5 (3.9-48.2) | -0.67 (-0.18; 0.350) | 17.7 | 5.7 | 10.63 (10.41-10.79) | 9.13 | 1.50 (1.25 to 1.72) | 0.28 (0.20-0.37) |
| Sédhiou | 14.4 (3.8-34.9) | -0.18 (-0.07; 0.757) | 12.8 | 9.9 | 10.85 (10.71-10.99) | 9.01 | 1.84 (1.66 to 2.01) | 1.57 (1.17-2.06) |
| Kaffrine | 12.5 (3.1-32.2) | -1.06 (-0.57; 0.002) | 5.2 | 11.6 | 10.44 (10.17-10.66) | 9.11 | 1.33 (1.04 to 1.58) | 1.40 (0.92-2.04) |
| Dakar | 11.0 (4.0-32.8) | -1.05 (-0.52; 0.005) | 5.8 | 6.1 | 10.30 (10.20-10.40) | 9.75 | 0.55 (0.35 to 0.75) | 2.68 (2.38-3.00) |
| Ziguinchor | 10.3 (3.8-17.6) | -0.50 (-0.35; 0.064) | 6.8 | 5.4 | 10.74 (10.57-10.87) | 9.04 | 1.70 (1.49 to 1.86) | 1.48 (1.04-2.05) |
| Fatick | 10.2 (1.0-28.1) | -1.39 (-0.50; 0.006) | 2.8 | 10.6 | 10.62 (10.41-10.88) | 9.17 | 1.45 (1.21 to 1.75) | 1.69 (0.97-2.75) |
| Thiès | 8.6 (2.2-23.8) | -1.07 (-0.52; 0.005) | 3.6 | 5.0 | 10.16 (10.04-10.28) | 9.30 | 0.85 (0.67 to 1.03) | 2.61 (2.13-3.17) |
| Matam | 7.6 (0.1-16.7) | -0.30 (-0.25; 0.195) | 9.1 | 8.3 | 9.62 (9.47-9.78) | 9.12 | 0.50 (0.30 to 0.71) | 1.48 (0.94-2.23) |
| Louga | 2.9 (1.3-5.7) | -0.09 (-0.25; 0.195) | 3.0 | 5.5 | 10.16 (9.98-10.38) | 9.22 | 0.94 (0.68 to 1.19) | 1.86 (1.16-2.81) |
| Saint-Louis | 1.7 (0.6-3.8) | -0.15 (-0.48; 0.009) | 1.3 | 4.4 | 10.35 (10.09-10.58) | 9.23 | 1.12 (0.82 to 1.41) | 7.00 (5.14-9.31) |
| **Senegal** | **24.5 (14.5-34.3)** | **+0.12 (0.08; 0.690)** | **19.3** | **11.7** | **10.14 (10.00-10.27)** | **9.08** | **1.06 (0.88 to 1.23)** | **0.72 (0.68-0.76)** |

*Incidence and centre of gravity of confirmed cases from the PNLP regional database (2009-2024); TPR, share of children under five and deaths from the district reports. Centre of gravity: circular mean month (9 = mid-September, 10 = mid-October). EIR: VECTRI driven by MRI-ESM2-0, 1995-2014. Lag: observed cases behind simulated EIR, with 95 % bootstrap CI. Trend: Mann-Kendall test and Sen slope on annual incidence, 2009-2024. Regions ordered by mean incidence.*

### Evaluation of the CMIP6 forcing

Over 1995-2014, the national mean annual rainfall of the ten bias-adjusted models ranged from 0.5 % (IPSL-CM6A-LR) to 23.2 % (EC-Earth3) above CHIRPS (638 mm), with MRI-ESM2-0 at +14.2 % (728 mm) (Fig 6). The correlation between the simulated and observed mean seasonal cycles was at least 0.99 for all models, and the rainfall centre of gravity differed from CHIRPS by 0.02 to 0.19 month nationally (MRI-ESM2-0: +0.07 month, about two days). Regionally, the bias of MRI-ESM2-0 ranged from +1 % (Ziguinchor) and +4 % (Kédougou) to +27 % (Kaolack, Dakar) and +29 % (Kaffrine), and the timing difference from -0.10 (Dakar) to +0.14 month (Saint-Louis). The forcing therefore reproduces the timing of the rainy season, with a wet bias of 10-30 % relative to CHIRPS in the centre and west.

**Fig 6.**
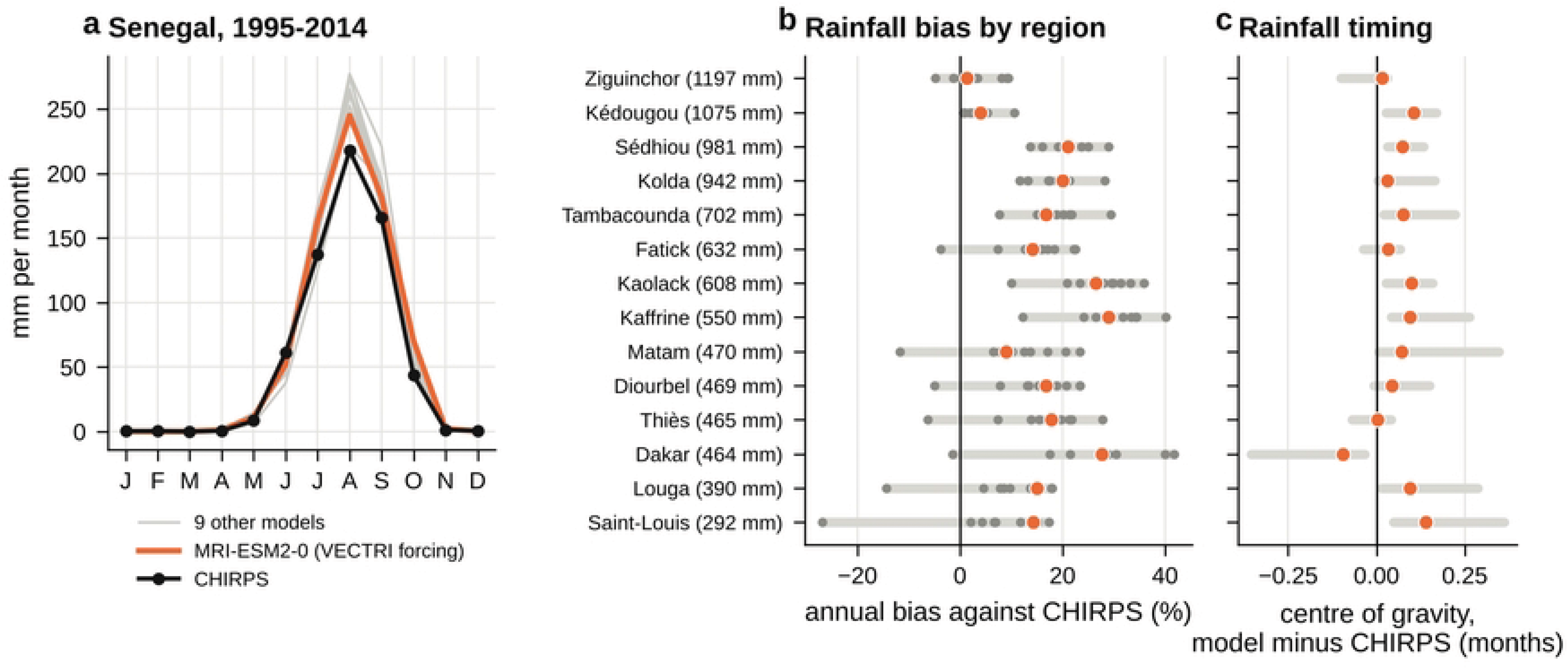
Evaluation of the NEX-GDDP-CMIP6 historical rainfall against CHIRPS, 1995-2014. (a) National mean monthly rainfall of CHIRPS, MRI-ESM2-0 (VECTRI forcing) and the nine other models. (b) Bias of annual rainfall by region (grey bars: range of the ten models; dots: individual models; orange: MRI-ESM2-0); CHIRPS annual rainfall in brackets. (c) Difference of rainfall centre of gravity, model minus CHIRPS (bars: range of the ten models).

### Transmission simulated by VECTRI

Driven by MRI-ESM2-0, VECTRI simulated a national mean annual EIR of 404 infective bites per person per year (10th-90th percentiles of years 312-518) over 1995-2014, with a single wave: 20.6 bites per person in July, 85.3 in August, 151.4 in September, 106.6 in October and 29.4 in November (Fig 7; S5 Fig). The EIR peaked in September in 16 of the 20 years and in October in four years; its centre of gravity was 9.08 (95 % CI 8.98-9.20), the concentration R was 0.85, 75 % of the annual EIR occurred within three months, and 90.7 % between July and October. The EIR exceeded 500 bites per person per year in Kolda (707), Sédhiou (656), Kédougou (583) and Tambacounda (520), and was low in Thiès (44) and negligible in Dakar (0.3), where the high population density reduces the number of bites per person in VECTRI (Fig 7a). The longest run of months with EIR ≥ 1 averaged 7.9 months, with the longest seasons in the centre and south-west (Fig 7b). The simulated timing varied little in space (centre of gravity 8.95-9.78 across departments; Fig 7c), and the detectable parasite ratio averaged 72 % (Fig 7d), a level that reflects the absence of calibration and of interventions in the simulations rather than the current prevalence. Simulated new cases had the same timing as the EIR (difference 0.02 month) and were highest where the EIR was moderate: 0.92 and 0.82 new cases per person per year in Thiès and Fatick against 0.29-0.37 in Kolda and Kédougou, a consequence of the acquired immunity represented by the model.

**Fig 7.**
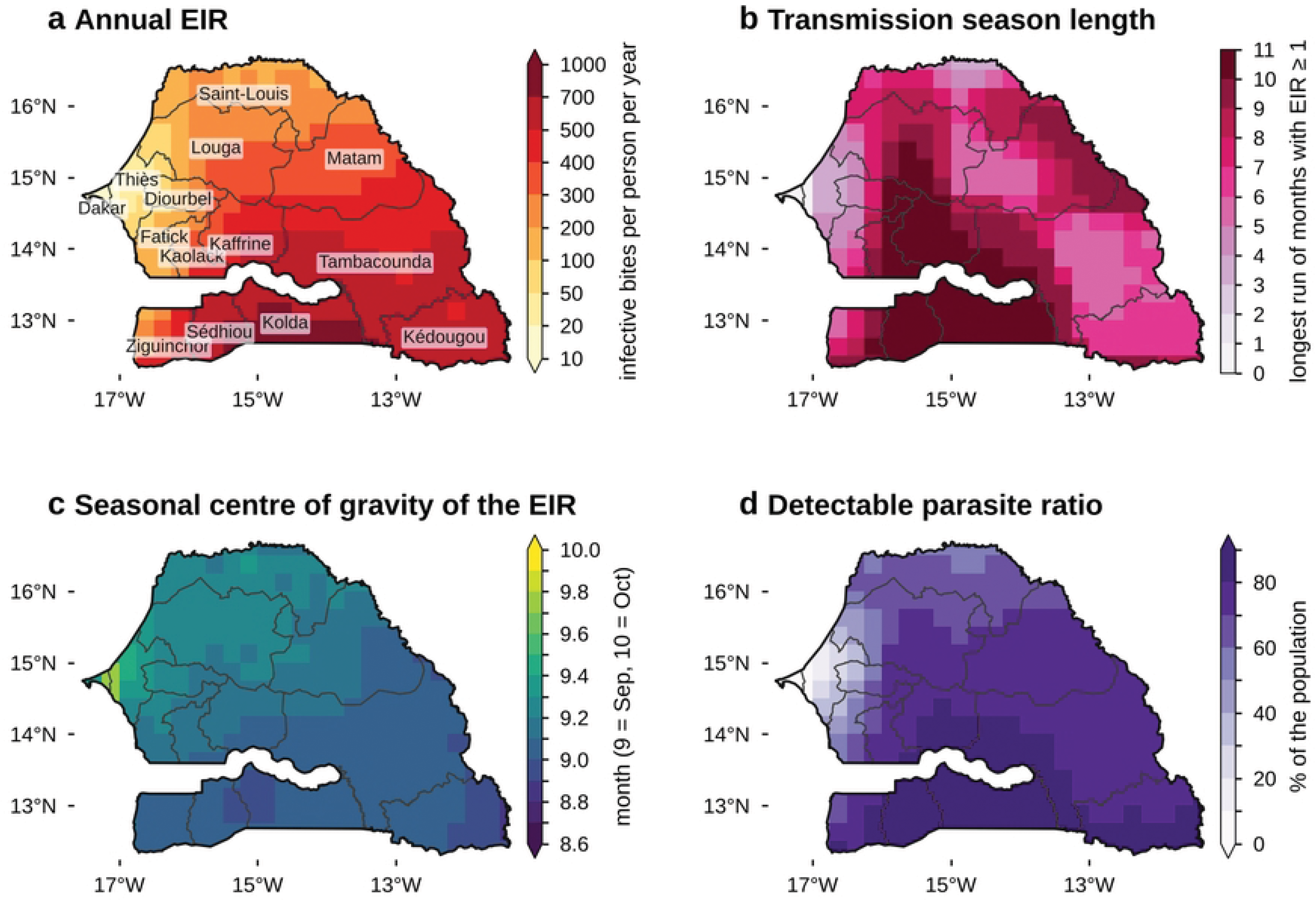
Transmission simulated by VECTRI driven by MRI-ESM2-0, 1995-2014. (a) Annual EIR. (b) Transmission season length (longest run of months with EIR ≥ 1 infective bite per person). (c) Seasonal centre of gravity of the EIR (circular mean month). (d) Detectable parasite ratio (uncalibrated model without interventions).

### Timing of observed cases and simulated transmission

Nationally, observed cases lagged the simulated EIR by 1.06 months (95 % CI 0.88-1.23) and the simulated new cases by 1.05 months (0.86-1.22) (Fig 8a; Table 2). The normalised cycles were most strongly correlated at a lag of one month (r = 0.97, against 0.66 without lag; Fig 8b); the cycle of the TPR (2016-2024) was also most correlated with the EIR at one month (r = 0.84), with a similar value at two months (0.83). The lag was robust to the observation period and source: 1.17 months (1.01-1.33) with the observations of 2009-2014, which overlap the simulated period, 1.13 (0.95-1.29) with 2009-2020, 1.11 (0.95-1.26) without 2022, and 0.99 (0.82-1.16) with the district reports of 2016-2024. Hospitalised cases lagged the EIR by 1.21 months (1.04-1.36), malaria deaths by 1.11 months (0.92-1.31) and tested patients, which include non-malarial fevers, by 0.70 month (0.51-0.89).

**Fig 8.**
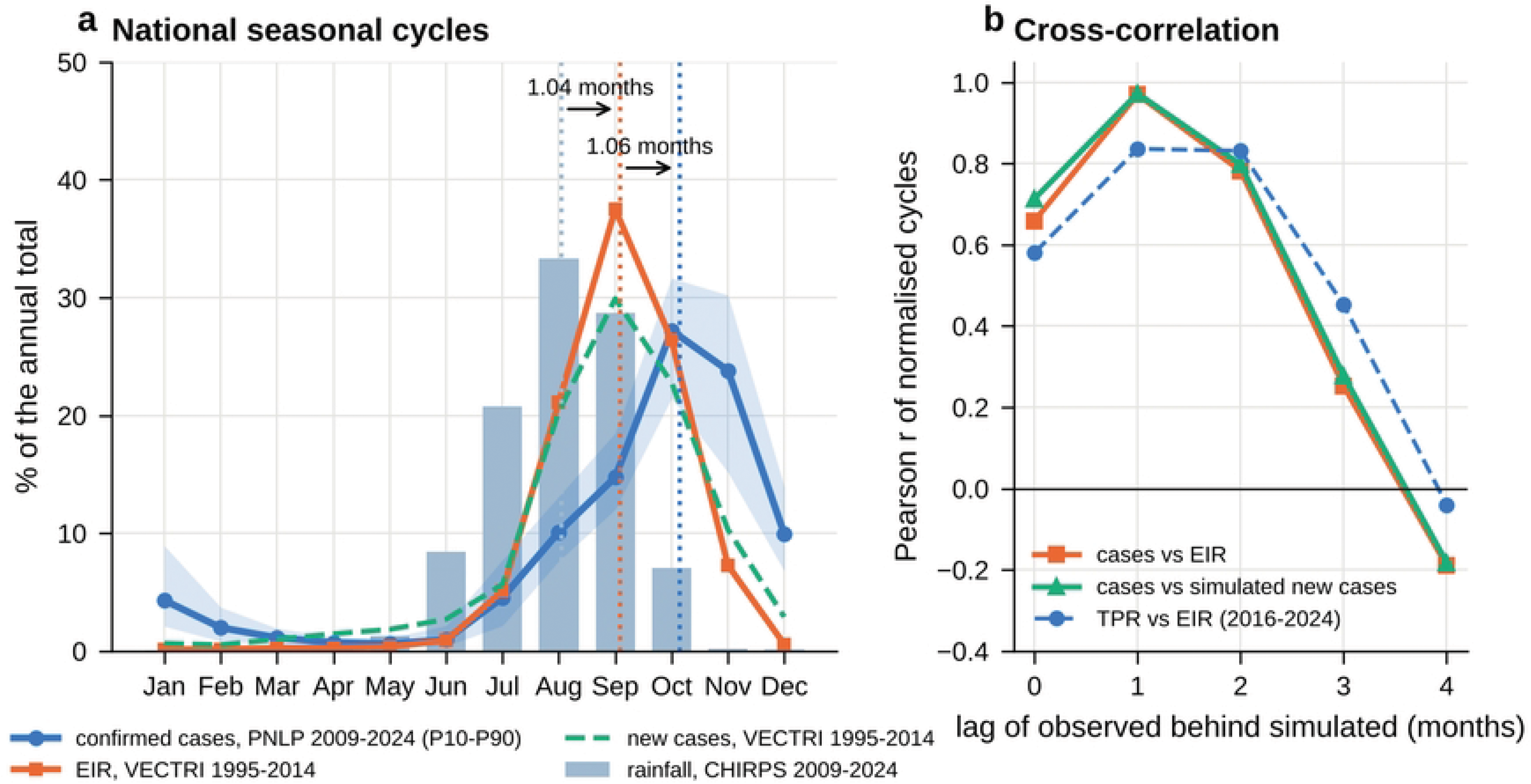
National seasonal cycles and their cross-correlation. (a) Monthly share of the annual total: confirmed cases (PNLP regional database, 2009-2024, with the 10th-90th percentile band of individual years), EIR and new cases simulated by VECTRI (1995-2014) and CHIRPS rainfall (bars, 2009-2024). Dotted lines: centres of gravity of rainfall, EIR and confirmed cases; arrows: delays in months. (b) Pearson correlation between the normalised mean cycles for lags of 0 to 4 months of the observed series behind the simulated series.

Observed cases followed CHIRPS rainfall (centre of gravity 8.04, mid-August) by 2.10 months (1.95-2.24) (Fig 8a). In VECTRI, the EIR followed the simulated rainfall by 1.04 months (0.89-1.19), and the timing of the forcing rainfall differed from CHIRPS by only 0.07 month, as did the CHIRPS timing between 1995-2014 and 2009-2024. The delay between rainfall and reported cases thus divides into two parts of about one month each: the delay between rainfall and transmission, reproduced by VECTRI, and the delay between infective bites and recorded cases (incubation, onset of symptoms, care seeking, diagnosis and reporting), which the model does not include.

Observed cases lagged the simulated EIR in all regions, with a CI excluding zero in 13 of 14 regions: from 0.50 month (95 % CI 0.30-0.71) in Matam, 0.55 in Dakar and 0.71 in Tambacounda to 1.69 in Diourbel, 1.70 in Ziguinchor and 1.84 (1.66-2.01) in Sédhiou; in Kédougou, the lag was 0.18 month (-0.02 to 0.38) (Fig 9; Table 2). The best cross-correlation lag was one month in ten regions, two months in Ziguinchor and Sédhiou and zero in Dakar and Kédougou, with maximum correlations of 0.89-0.98 (S6 Fig, S7 Fig). The delay between simulated rainfall and EIR was nearly the same in all regions (0.99-1.08 month, except Dakar, 1.57), whereas the delay between observed rainfall and cases ranged from 1.29 (Kédougou) to 2.82 months (Sédhiou): the regional differences in the observed lag arise after transmission, not in the timing of rainfall. Across departments, the lag ranged from -0.16 to 1.88 months (median 1.04), with a CI above zero in 44 of 45 departments; it was shortest in the east and north-east and longest in the centre-west (Kaolack, Fatick, Diourbel) and in Casamance (Fig 10c). The simulated timing did not reproduce the spatial variations of the observed timing (Spearman ρ = 0.11, p = 0.70, across regions; ρ = 0.06, p = 0.69, across departments), whereas the observed timing was weakly related to the timing of observed rainfall across departments (ρ = 0.33, 95 % CI 0.00 to 0.59, p = 0.028).

**Fig 9.**
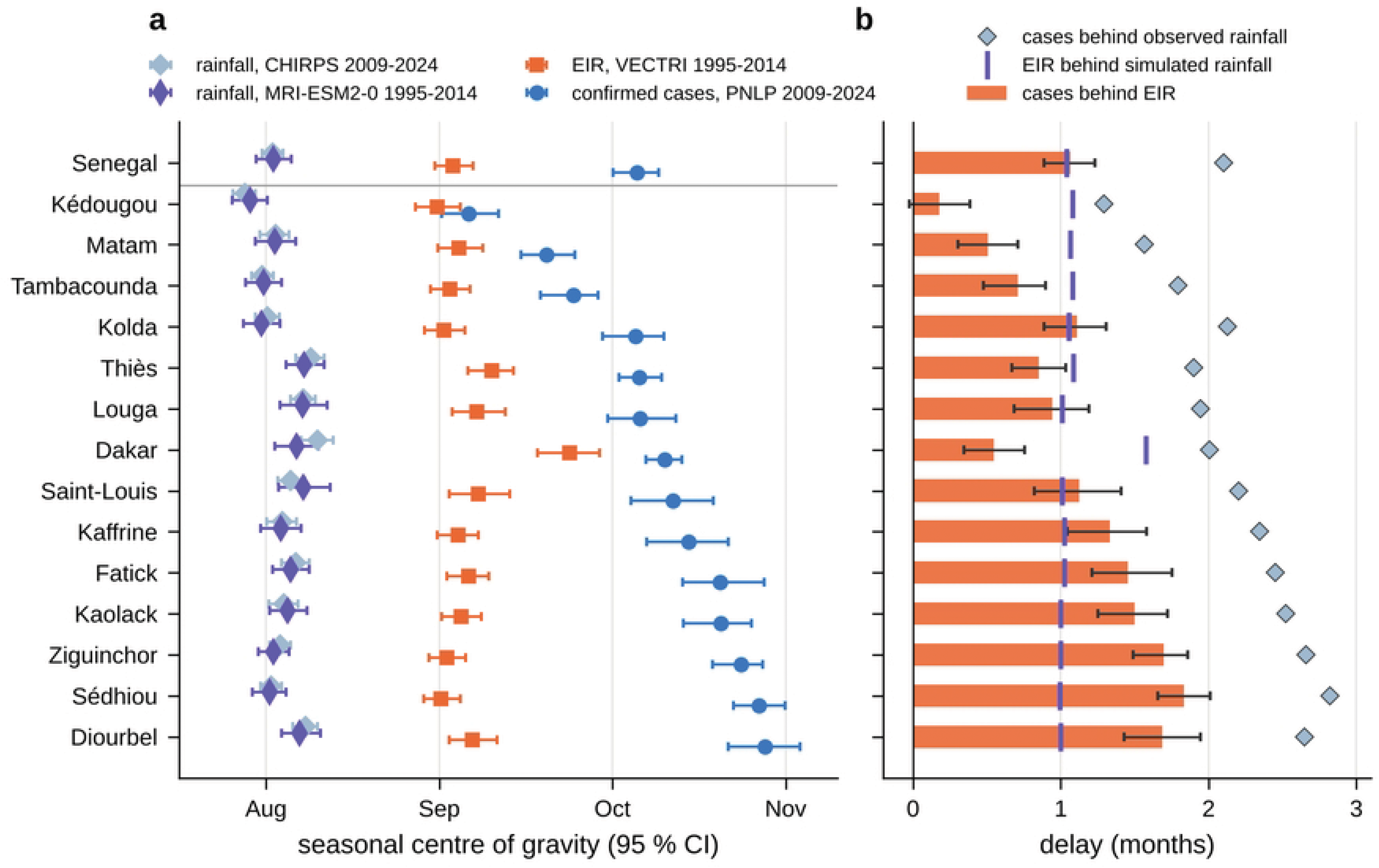
Timing of rainfall, simulated transmission and confirmed cases by region. (a) Seasonal centre of gravity with 95 % bootstrap CI of CHIRPS rainfall (2009-2024), MRI-ESM2-0 rainfall and VECTRI EIR (1995-2014) and confirmed cases (2009-2024). (b) Delays: cases behind EIR (bars, 95 % CI), cases behind observed rainfall (diamonds) and EIR behind simulated rainfall (vertical marks). Regions ordered by the timing of confirmed cases.

**Fig 10.**
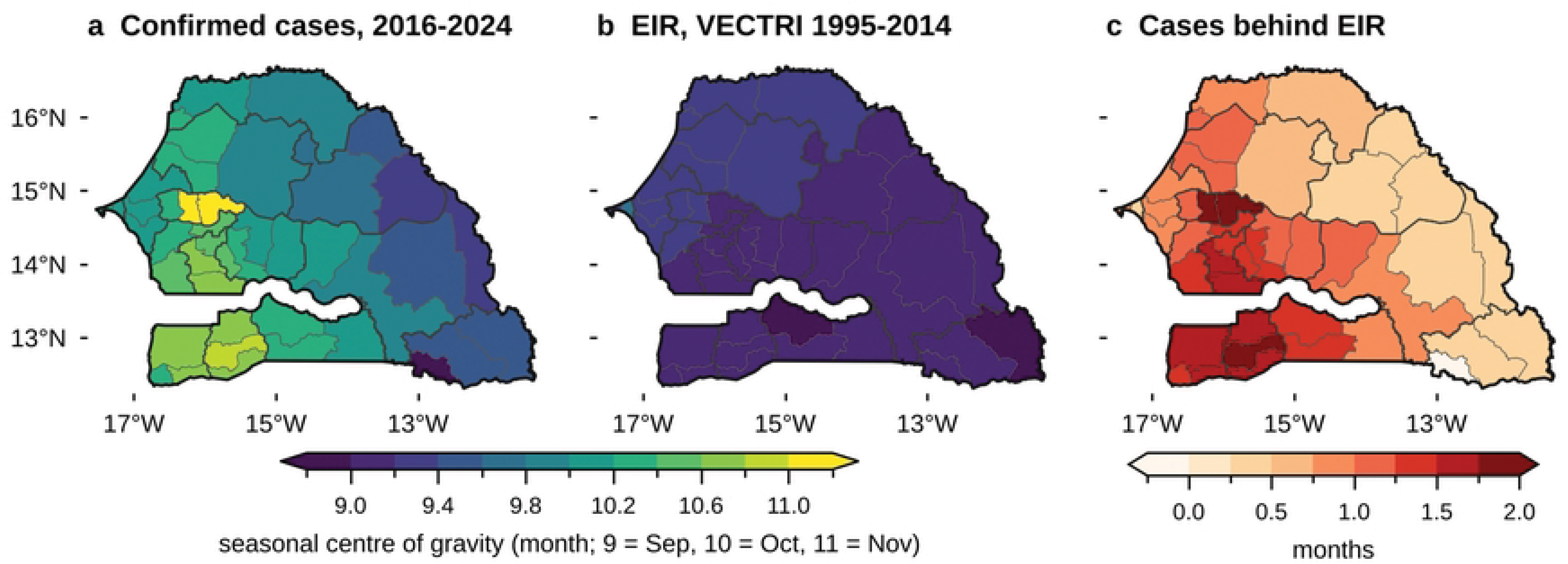
Seasonal timing by department. Centre of gravity of (a) confirmed cases (district reports, 2016-2024) and (b) EIR simulated by VECTRI (1995-2014); (c) lag of confirmed cases behind the simulated EIR.

### Spatial pattern of transmission intensity

The simulated EIR reproduced the gradient of transmission intensity. Across the 45 departments, the TPR over 2016-2024 was correlated with the simulated annual EIR (Spearman ρ = 0.67, 95 % CI 0.47-0.79) and detectable parasite ratio (ρ = 0.63, 0.45-0.75), more strongly than with the annual rainfall of CHIRPS (ρ = 0.59, 0.31-0.78) (Fig 11a). Across regions, the simulated EIR was correlated with incidence (ρ = 0.56, 0.08-0.86, p = 0.037) and with the TPR (ρ = 0.64, p = 0.013) (Fig 11c). In contrast, simulated new cases per person were inversely correlated with the TPR across departments (ρ = -0.55, -0.73 to -0.31) (Fig 11b), and the regional distribution of cases obtained by multiplying simulated new cases by the AfriPop population did not match the observed distribution (ρ = -0.29, -0.65 to 0.24): VECTRI placed 33.1 % of the national cases in Thiès, 15.9 % in Fatick and 14.3 % in Louga (observed 4.9 %, 2.4 % and 0.8 %), and 1.6 %, 0.7 % and 2.6 % in Kolda, Kédougou and Tambacounda (observed 22.0 %, 13.8 % and 19.0 %).

**Fig 11.**
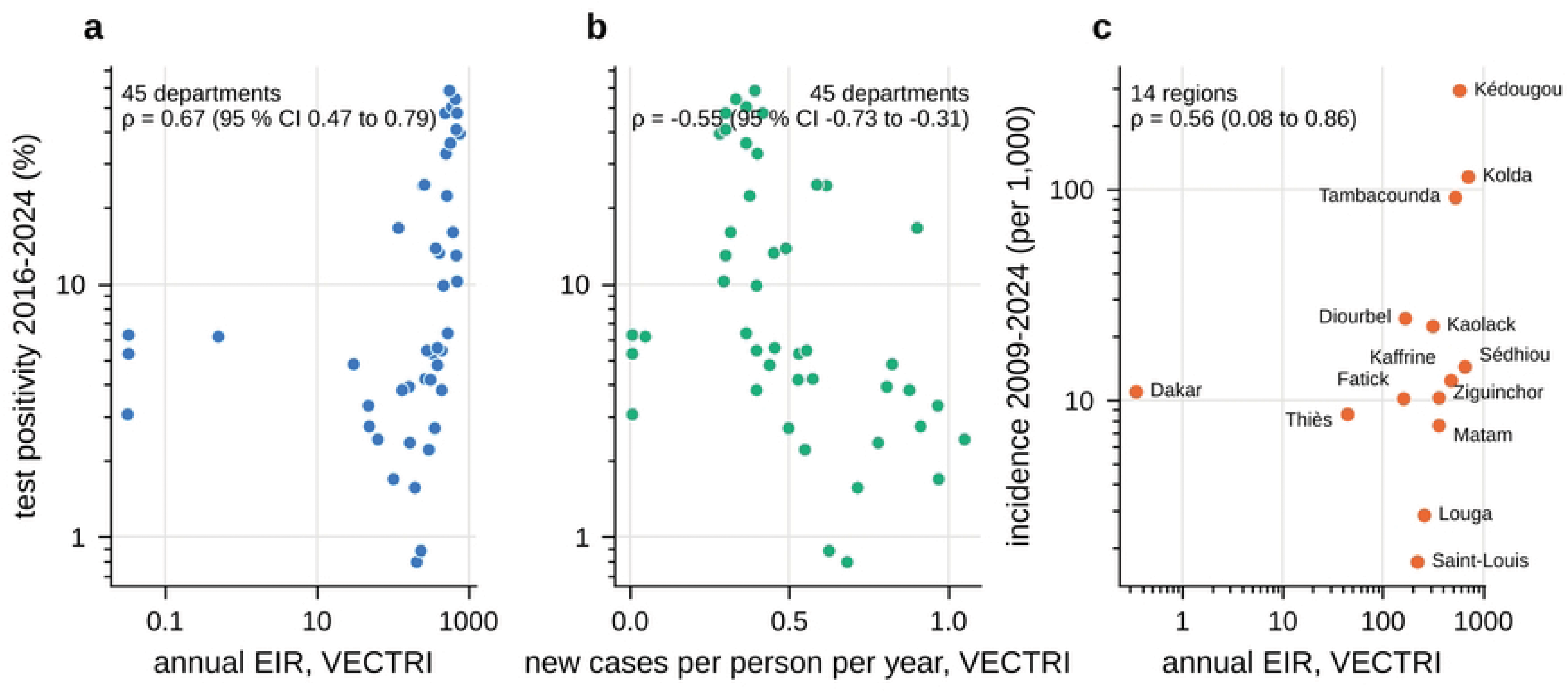
Spatial consistency between simulated transmission and observed malaria. (a) Test positivity rate (2016-2024) against the simulated annual EIR and (b) against simulated new cases per person, 45 departments. (c) Mean incidence (2009-2024) against the simulated annual EIR, 14 regions. Spearman correlations with 95 % bootstrap CI over units; log scales.

### Severity

Hospitalised malaria cases ranged from 9,179 to 15,700 per year, i.e. 2.1-3.4 % of confirmed cases in 2016-2024 and 4.8 % in 2025 (Table 1). Malaria deaths reported in DHIS2 decreased every year from 399 in 2021 to 274 in 2024 and 155 in 2025 (τ = -1.00, p = 0.017, n = 5; Sen slope -55 deaths per year, 95 % CI -119 to -23) (Fig 12a). Over 2021-2024, deaths per 1,000 confirmed cases were lowest in the high-transmission regions (Kaolack 0.28, 95 % CI 0.20-0.37; Kédougou 0.31, 0.25-0.37; Kolda 0.40, 0.34-0.46; Tambacounda 0.50, 0.43-0.58) and highest in Saint-Louis (7.00, 5.14-9.31), Dakar (2.68, 2.38-3.00) and Thiès (2.61, 2.13-3.17) (Fig 12b).

**Fig 12.**
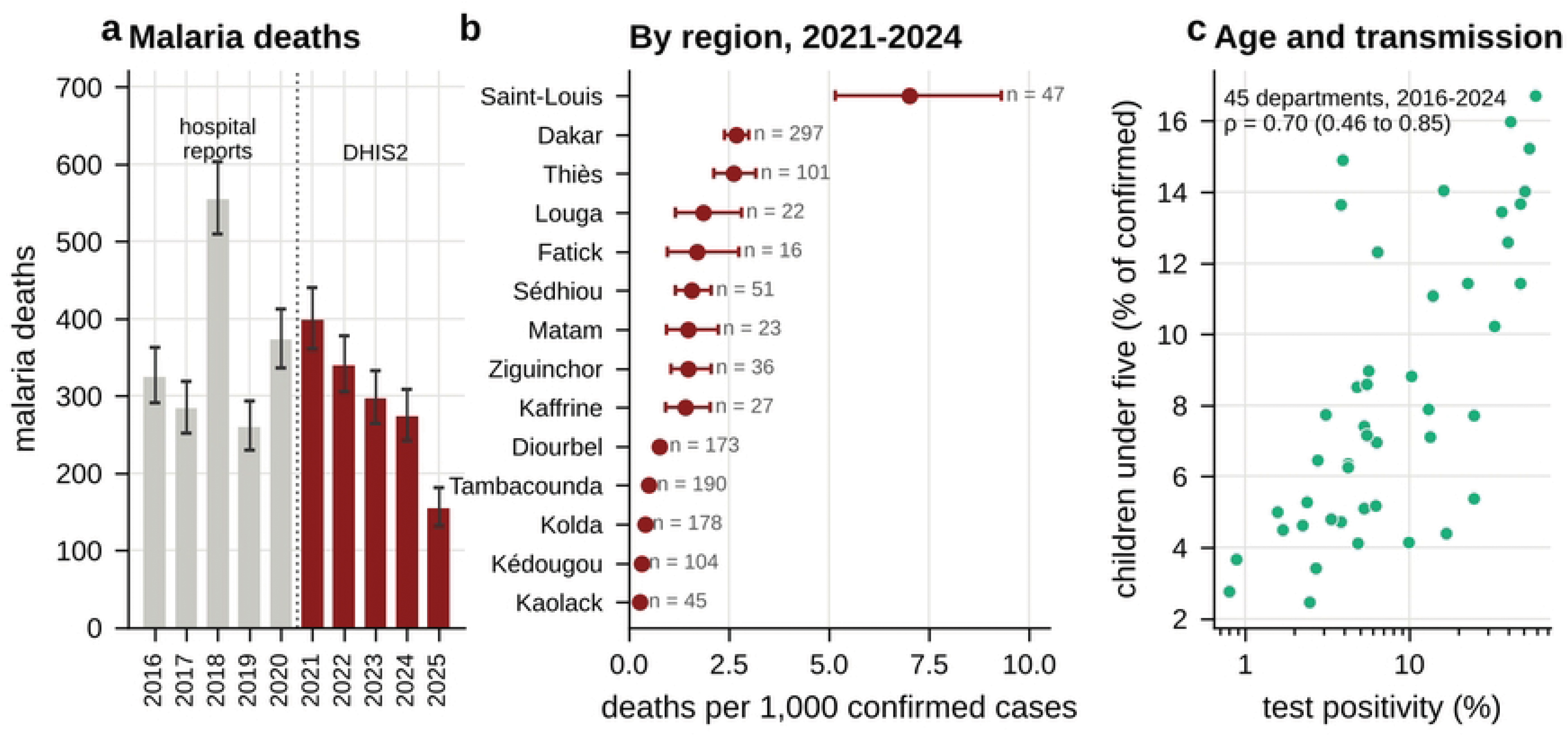
Severity and age. (a) Malaria deaths with exact Poisson 95 % CI: hospital reports (2016-2020, grey) and DHIS2 (2021-2025, red; 2025 provisional). (b) Deaths per 1,000 confirmed cases by region, 2021-2024, with exact Poisson 95 % CI (n: number of deaths); the ratio is not a case fatality rate. (c) Share of children under five among confirmed cases against the test positivity rate, 45 departments, 2016-2024.

### The 2025 data and the LLIN mass campaign

In 2025, 3.95 million LLINs were distributed in 34 districts covering all districts of Kaffrine, Kaolack, Kolda, Kédougou, Matam, Sédhiou, Tambacounda and Ziguinchor and one district of Fatick (Sokone) (Fig 13a). Confirmed cases decreased by 55.7 % in these districts relative to the mean of 2021-2024 (from 354,512 to 157,077) and by 9.7 % in the other 45 districts (from 99,817 to 90,102). At the district level, the median ratio of 2025 to 2021-2024 was 0.60 (interquartile range 0.30-0.86) for confirmed cases in campaign districts against 0.80 (0.66-1.10) elsewhere (Mann-Whitney p = 0.003), whereas the number of tested patients was unchanged in campaign districts (median ratio 1.00) and increased elsewhere (1.31; p < 0.001); as a result, the TPR ratio was similar (0.58 and 0.59; p = 0.081) (Fig 13b). In campaign districts, monthly confirmed cases were close to or above the 2021-2024 level from January to June 2025 (ratios 0.88-1.55) and fell from July (0.74) to November (0.23), while the number of tested patients also decreased in October-December (ratios 0.62-0.73) (Fig 13c). The national TPR fell to 9.1 % in 2025. Kolda, where 2025 incidence was 0.13 times the 2016-2024 median, was flagged by the PNLP as possibly incomplete.

**Fig 13.**
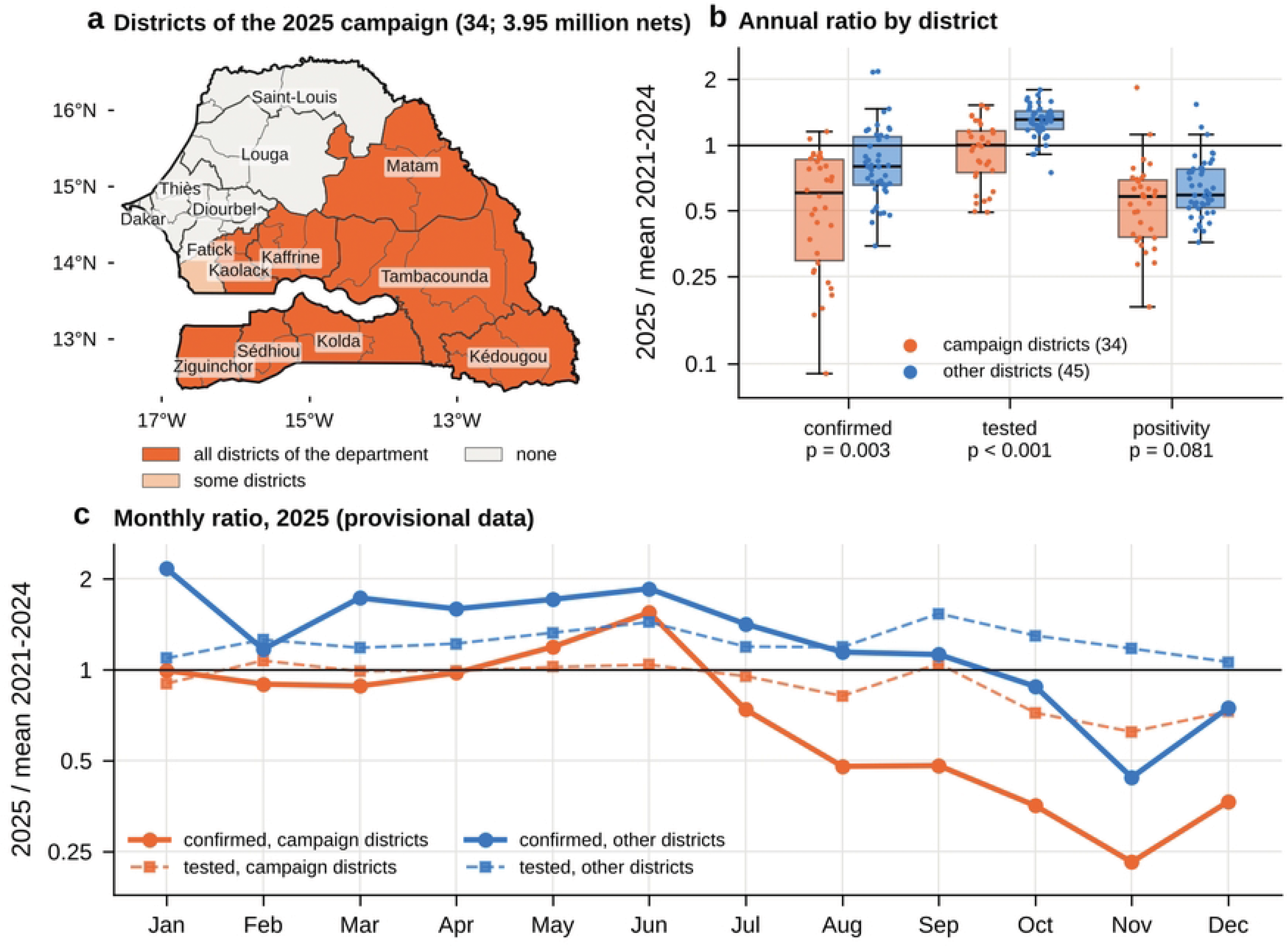
Confirmed malaria in 2025 and the LLIN mass campaign. (a) Departments with all, some or none of their districts covered by the 2025 campaign (34 districts; 3.95 million nets distributed). (b) Ratio of 2025 to the mean of 2021-2024 by district for confirmed cases, tested patients and test positivity, campaign districts (orange) and other districts (blue), with Mann-Whitney p values. (c) Monthly ratio of 2025 to the 2021-2024 mean. 2025 data are provisional.

### Projected changes in climate

Over Senegal, the ten models projected warming of +1.4 °C (range 1.0 to 2.1) in 2041-2060 and +2.3 °C (1.5 to 3.4) in 2081-2100 under SSP2-4.5, and +1.9 °C (1.4 to 2.4) and +4.6 °C (2.9 to 5.8) under SSP5-8.5 (Fig 14a; Table 3). The median change in annual rainfall was negative in all cases (-8 %, -13 %, -13 % and -32 %), but the models disagreed: two or three of the ten models, mainly EC-Earth3 and INM-CM5-0, projected wetter conditions (up to +25 %) (Fig 14b, 14c; S8 Fig). Under SSP5-8.5 in 2081-2100, the decrease exceeded 30 % in most regions of the centre, east and south (-35 % in Kaffrine and -34 % in Matam) and was smaller near the coast (-19 % in Dakar and Thiès), and at least eight of ten models agreed on a decrease over most of the country (Fig 14e). The decrease concerned June to September, with a median of -98 mm in August under SSP5-8.5 in 2081-2100, whereas October rainfall increased slightly (+6 to +16 mm), so that the rainfall centre of gravity moved later by 0.07 to 0.22 month (Fig 14f). MRI-ESM2-0 was close to the median for temperature and slightly drier for rainfall (-9 %, -19 %, -12 % and -44 %).

**Fig 14.**
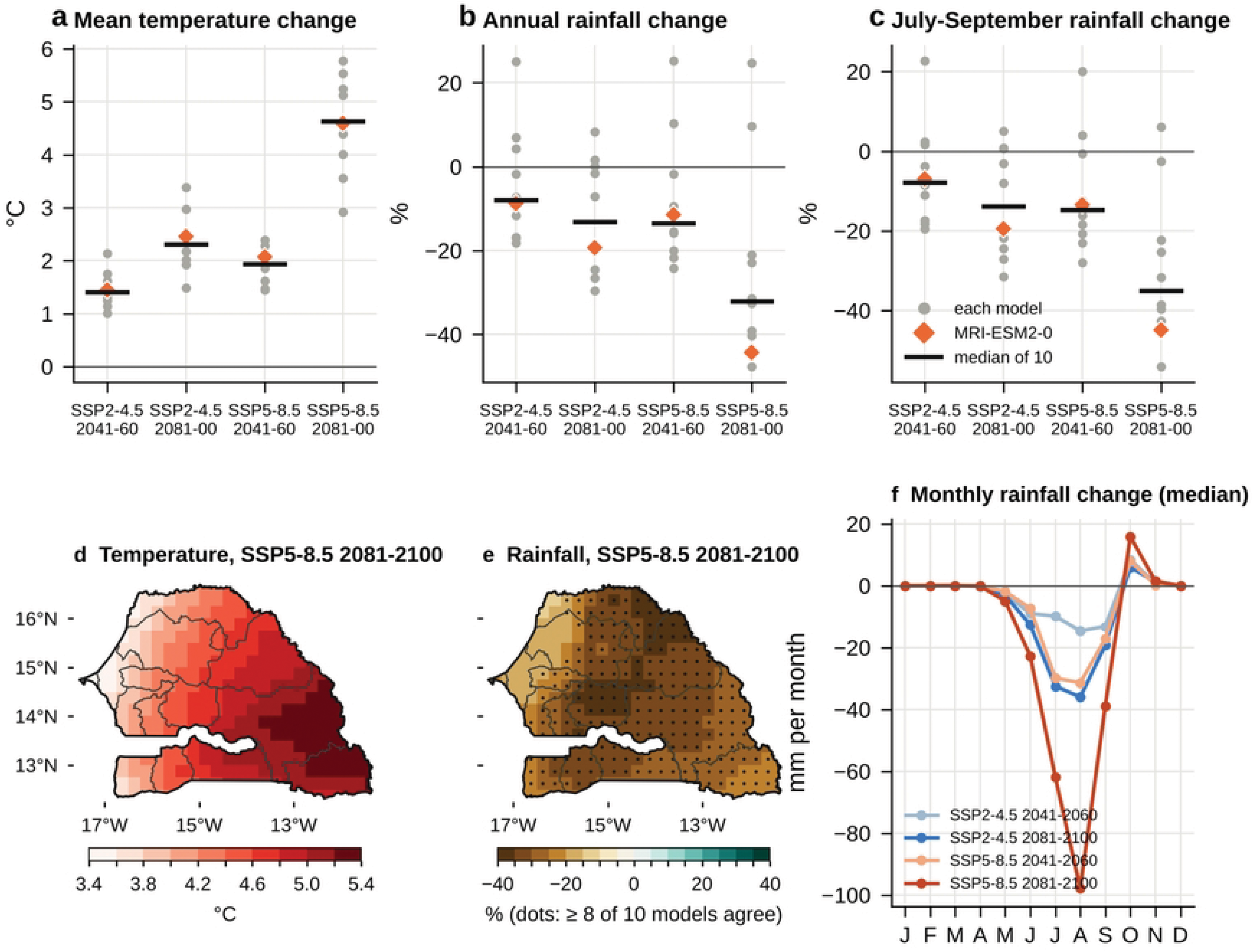
Projected changes in temperature and rainfall over Senegal, ten NEX-GDDP-CMIP6 models, relative to 1995-2014. (a) Mean temperature, (b) annual rainfall and (c) July-September rainfall: individual models (grey), MRI-ESM2-0 (orange) and median (black bar). (d) Median change in temperature and (e) in annual rainfall under SSP5-8.5 in 2081-2100 (dots: at least 8 of 10 models agree on the sign). (f) Median change in monthly rainfall.

**Table 3.** Projected changes over Senegal relative to 1995-2014.

| **Indicator** | **1995-2014** | **SSP2-4.5 2041-2060** | **SSP2-4.5 2081-2100** | **SSP5-8.5 2041-2060** | **SSP5-8.5 2081-2100** |
|---|---|---|---|---|---|
| Mean temperature (°C), 10 models: median (range) | 28.5 | +1.4 (1.0 to 2.1) | +2.3 (1.5 to 3.4) | +1.9 (1.4 to 2.4) | +4.6 (2.9 to 5.8) |
| Annual rainfall (%), 10 models: median (range); models wetter | 727 mm | -8 (-18 to +25); 3 | -13 (-30 to +8); 3 | -13 (-24 to +25); 2 | -32 (-48 to +25); 2 |
| July-September rainfall (%), median |  | -8 | -14 | -15 | -35 |
| Suitable season, rules (months), median; models with significant change | 3.4 | -0.12; 2 | -0.34; 6 | -0.29; 3 | -0.98; 9 |
| Months above 34 °C, median | 0.3 | +0.7 | +1.2 | +0.9 | +3.6 |
| Area with stable suitability (%) | 64 | 61 | 57 | 60 | 33 |
| Rainfall, MRI-ESM2-0 (%) | 728 mm | -9 | -19 | -12 | -44 |
| Annual EIR, VECTRI (bites per person) | 404 | +6 % (p = 0.90) | +3 % (p = 0.52) | +9 % (p = 0.27) | -35 % (p < 0.001) |
| Transmission season, VECTRI (months) | 7.9 | +0.11 (p = 0.40) | +0.29 (p = 0.07) | +0.49 (p = 0.005) | -0.28 (p = 0.16) |
| EIR centre of gravity, VECTRI (months; 95 % CI) | 9.08 | +0.07 (-0.07 to 0.20) | -0.06 (-0.21 to 0.08) | +0.05 (-0.12 to 0.21) | +0.06 (-0.10 to 0.22) |
| Share of annual EIR in July-October (%) | 90.7 | +1.1 points | +0.8 points | -0.9 points | -0.8 points |
| Detectable parasite ratio, VECTRI (%) | 72.2 | -0.4 points | -1.9 points | +0.4 points | -16.1 points |

*Climate: ten NEX-GDDP-CMIP6 models (median and range of the changes; number of models projecting wetter conditions). Suitability: rule-based season (16-34 °C, ≥ 60 mm), median of ten models and number of models with a significant change (Mann-Whitney p < 0.05). VECTRI: driven by MRI-ESM2-0; p values from Mann-Whitney tests on annual values (20 years); centre of gravity change with 95 % bootstrap CI.*

### Projected changes in suitability and simulated transmission

With the rule-based approach, the national suitable season of 3.4 months (1995-2014), stable over 64 % of the area, changed little by mid-century (-0.12 and -0.29 month; significant in two and three models), and decreased by 0.34 month under SSP2-4.5 in 2081-2100 (six models significant) and by 0.98 month under SSP5-8.5 in 2081-2100 (nine models significant), when the stable area fell to 33 % and the number of months above 34 °C increased by 3.6 (Fig 15a; Table 3; S9 Fig). The largest decreases occurred in the east (Matam -1.7, Tambacounda -1.3, Kédougou -1.0 months under SSP5-8.5 in 2081-2100), as more months exceed 34 °C and rainfall decreases.

**Fig 15.**
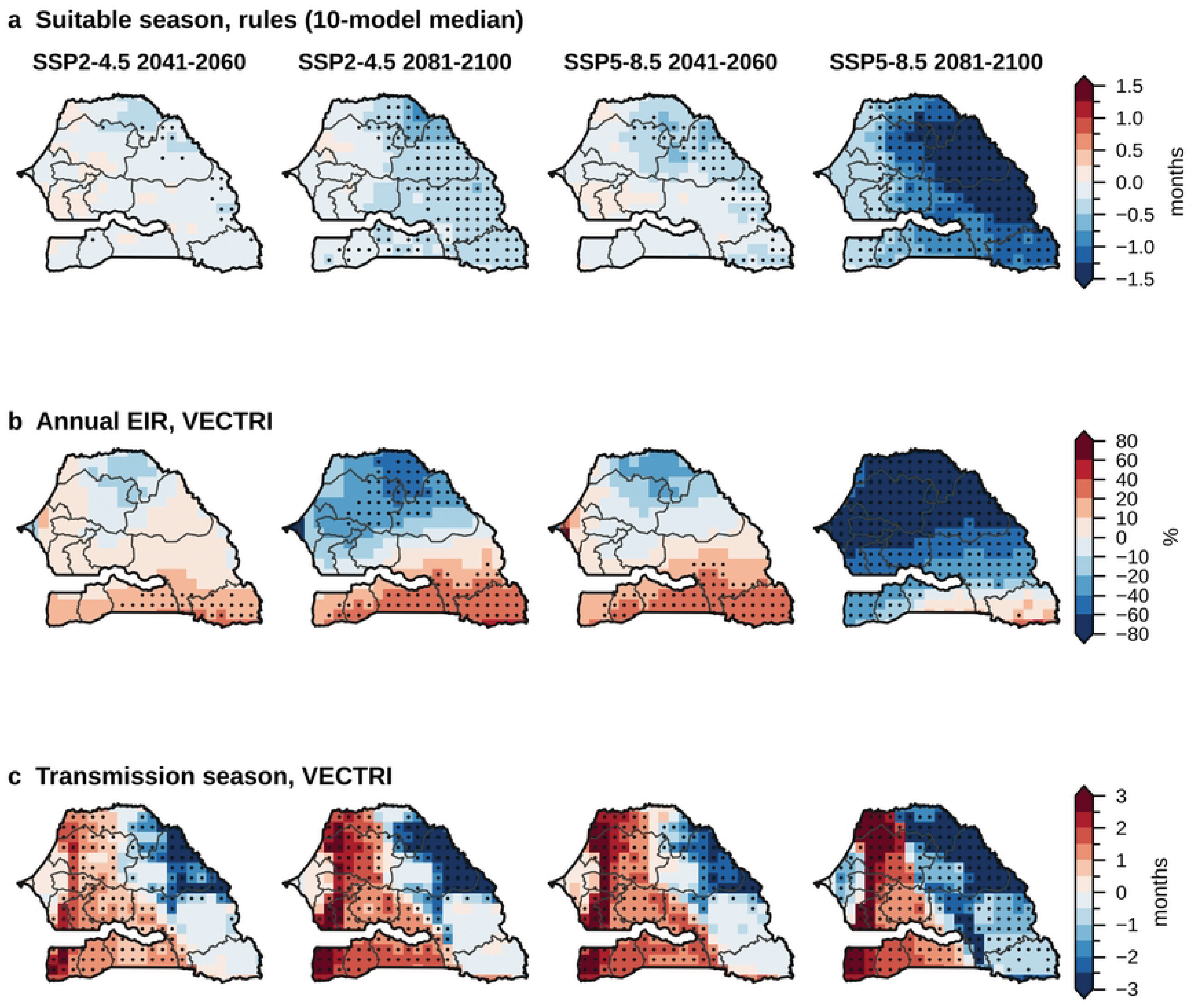
Projected changes in climatic suitability and simulated transmission, relative to 1995-2014. (a) Suitable season (rules, median of ten models; dots: at least 8 of 10 models agree on the sign). (b) Annual EIR and (c) transmission season length simulated by VECTRI driven by MRI-ESM2-0 (dots: Mann-Whitney p < 0.05 on annual values).

VECTRI driven by MRI-ESM2-0 simulated small and non-significant changes in national annual EIR under SSP2-4.5 (+6 % in 2041-2060, +3 % in 2081-2100) and SSP5-8.5 in 2041-2060 (+9 %), but a decrease of 35 % under SSP5-8.5 in 2081-2100 (p < 0.001), together with a decrease of 16 percentage points of the parasite ratio (Fig 15b, Fig 16; Table 3). The changes had opposite signs in space: the EIR increased in the south and south-east under SSP2-4.5 and by mid-century (significantly in Kédougou and Kolda in all three cases, by 14 % to 32 %) and decreased in the north and west (-30 % in Louga and -37 % in Saint-Louis under SSP2-4.5 in 2081-2100). Under SSP5-8.5 in 2081-2100, the EIR decreased by 53 % to 90 % in the centre, north and west, by 19 % to 32 % in Sédhiou, Ziguinchor and Tambacounda, and did not change significantly in Kédougou (+7 %) and Kolda (-4 %). The transmission season lengthened in the west and south-west and shortened in the north-east (Fig 15c), with national changes between -0.28 and +0.49 month. The timing of simulated transmission did not change: the national EIR centre of gravity moved by -0.06 to +0.07 month, with CIs including zero in all four cases, and the share of the annual EIR falling in July-October remained between 89.8 % and 91.9 % (90.7 % in 1995-2014) (Fig 16).

**Fig 16.**
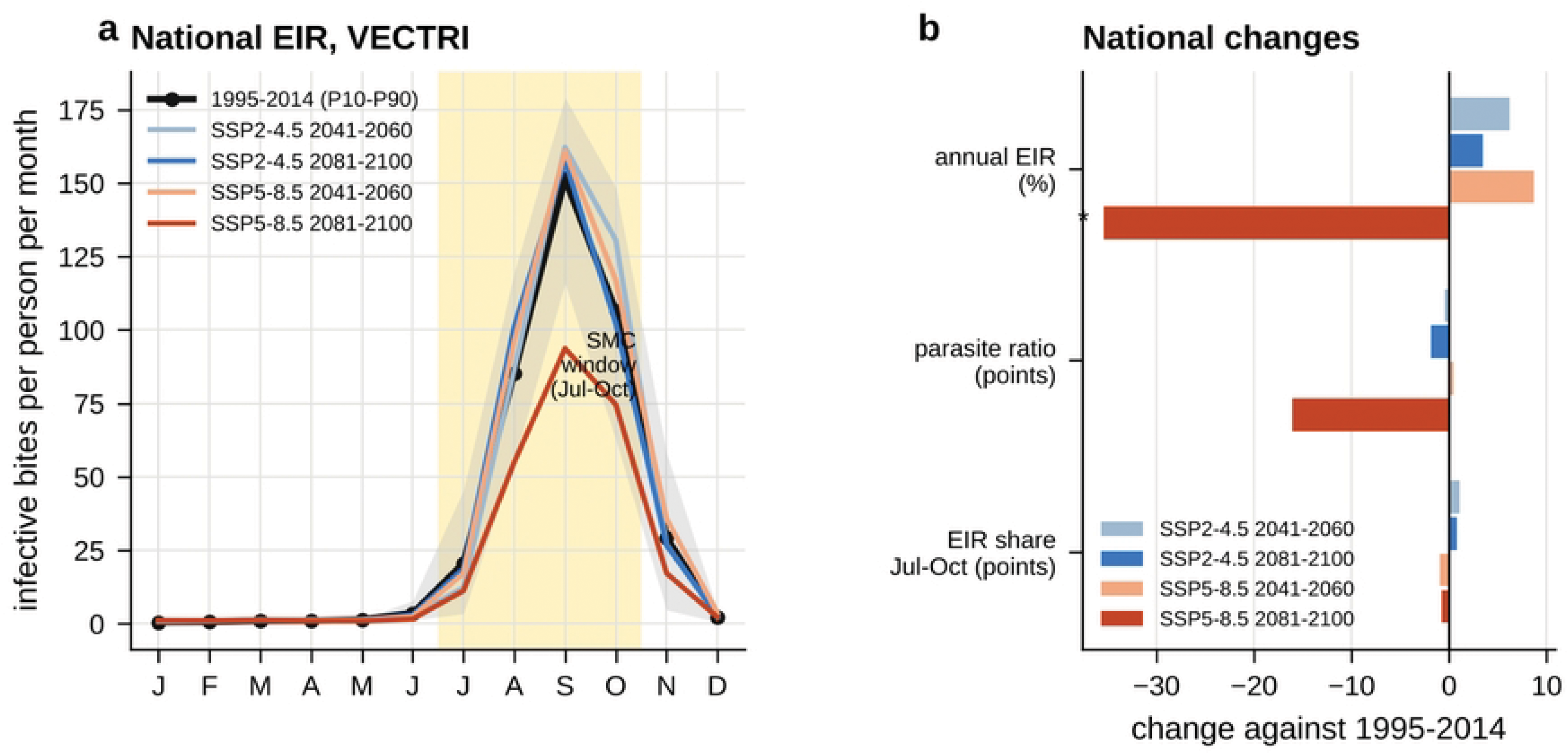
National EIR simulated by VECTRI in the future. (a) Mean monthly EIR for 1995-2014 (with 10th-90th percentiles of years) and for the four scenario-periods; shading: SMC window (July-October). (b) National changes in annual EIR, detectable parasite ratio and share of the EIR in July-October; *: Mann-Whitney p < 0.05.

## Discussion

### Principal findings

This study combines sixteen years of monthly regional surveillance, nine years of harmonised district reports with testing data, observed rainfall, ten bias-adjusted CMIP6 models and VECTRI simulations to evaluate the description of malaria seasonality in Senegal by a dynamical model and to project its changes. Four results stand out. First, the timing of confirmed malaria is regular, centred on mid-October, and reported cases lag the transmission simulated by VECTRI by about one month nationally and in 13 of 14 regions and 44 of 45 departments; this lag is robust to the observation period and source. Second, the delay between rainfall and reported cases (about two months) is shared equally between the delay between rainfall and transmission, which VECTRI reproduces with a nearly uniform value of one month, and the delay between transmission and recorded cases; the regional differences in observed timing arise in the second part and are not reproduced by the model. Third, the simulated EIR reproduces the spatial gradient of transmission intensity measured by the TPR, whereas the simulated cases do not reproduce the distribution of the burden. Fourth, the projected changes in timing are small, while the intensity of transmission is projected to decrease strongly only under SSP5-8.5 at the end of the century, with increases in the south-east under the other scenarios.

### Timing of transmission and reported cases

The one-month lag between simulated transmission and confirmed cases is consistent with previous modelling studies in Senegal, which placed the transmission peak in September-October and reported cases one to two months after the rainfall peak [2,3,13], and with entomological and epidemiological observations in the Sahel, where vector abundance precedes clinical malaria by several weeks [42]. It is compatible with an intrinsic incubation period of the order of two weeks for *Plasmodium falciparum* and with the delays of care seeking, diagnosis and reporting. Hospitalised cases followed outpatient cases by about one week, as expected for the progression to severe disease. With three quarters of annual cases within four consecutive months, malaria in Senegal meets the criterion of marked seasonality proposed by Roca-Feltrer et al. [43], which underlies eligibility for SMC [44].

The separation of the delay into two parts of about one month each is, to our knowledge, new for Senegal. It was made possible because the bias-adjusted forcing reproduces the timing of observed rainfall to within a few days, so that the simulated delay between rainfall and transmission can be compared with the observed delay between rainfall and cases. The uniformity of the simulated delay across regions contrasts with observed delays ranging from 1.3 months in Kédougou to 2.8 months in Sédhiou. Three types of factors not represented in VECTRI may explain this contrast. Hydrology: in Casamance and in the centre-west (Kaolack, Fatick, Diourbel), flooded areas, rice fields, the Saloum and Casamance estuaries and other persistent water bodies can maintain breeding sites after the end of the rains, as shown by pond-resolving hydrological modelling [11,42,45], which would extend transmission into November-December. Interventions: SMC delivered from July to October [6,46] reduces cases among children during the rounds and may shift the remaining cases later; the decrease of the share of children under five among confirmed cases over the period is consistent with this effect, but the data do not allow it to be quantified. Reporting and care seeking may also differ between regions. Conversely, the early season in Kédougou, where cases nearly coincide with simulated transmission, coincides with the earliest and most abundant rainfall and the highest burden, and suggests that the early part of the season is underestimated by the model in the south-east, where the season has also become later between 2009-2014 and 2015-2024.

### Spatial pattern

The TPR is well suited to evaluate dynamical malaria models with routine data. Because almost all suspected cases were tested, it measures the share of febrile patients with a malaria infection and does not depend on population estimates, which are uncertain at fine scale. Across departments, the simulated EIR was slightly more strongly correlated with the TPR than observed rainfall, although the confidence intervals overlap; the combination of rainfall, temperature and population represented by VECTRI therefore reproduces the gradient at least as well as rainfall alone. The concentration of the burden in Kédougou, Kolda and Tambacounda, where incidence increased over 2009-2024, is consistent with the persistent south-eastern hotspots identified over 2000-2020 [47] and with the gradient of simulated transmission [3,4,19,20]. In contrast, the negative association between simulated new cases and observed malaria reflects the representation of acquired immunity in VECTRI, which reduces the number of clinical cases where transmission is intense [10], and the AfriPop population at 1°, which spreads the population of Dakar and Thiès over neighbouring cells. In VECTRI, high population densities also dilute bites [10], which explains the near-zero EIR of Dakar, whereas the cases reported in Dakar may include infections acquired elsewhere and treated in the capital. The simulated case counts should therefore not be used to allocate resources between regions without calibration.

The higher share of children under five where the TPR is high, and the lower ratio of deaths to confirmed cases in high-transmission regions, are consistent with the age shift of clinical malaria with transmission intensity [48] and with the relation between transmission intensity and the clinical spectrum of severe malaria [49]. The high ratios in Saint-Louis, Dakar and Thiès may also reflect the referral of severe patients to regional and national hospitals and the lower immunity of populations with little exposure; patient-level data would be needed to distinguish these explanations.

### Surveillance indicators and the 2025 data

National incidence showed no trend over 2009-2024, but the TPR decreased significantly over 2016-2024 while testing remained near universal, and incidence increased in the south-east and decreased in the centre and west. A decreasing TPR with a stable number of confirmed cases indicates that more patients with non-malarial fevers were tested (1.56 million suspected cases in 2016, 2.91 million in 2024); national incidence alone would therefore hide both a change in testing practice and a geographical concentration of transmission. In 2025, confirmed cases decreased by more than half in the districts of the LLIN mass campaign, against less than 10 % elsewhere, with a stable number of tests in these districts and a decrease that started in July, after the first half of the year was close to the usual level. This pattern is consistent with an effect of the campaign on transmission, in line with the large contribution of insecticide-treated nets to the decrease of malaria in Africa [50]. It cannot, however, be attributed to the campaign from these data: the campaign districts were selected for their high burden, the 2025 data are provisional, the number of tests also decreased in October-December in the campaign districts, Kolda was flagged as possibly incomplete, and the decrease of confirmed cases in November in the other districts suggests incomplete late-year reporting. A verification of the 2025 data with the PNLP and a comparison with SMC coverage and rainfall will be needed.

### Projections

The ten models agree on warming but not on the sign of rainfall change, a known feature of CMIP projections over the Sahel [51]; the median decrease of annual rainfall concerns the core of the rainy season, while October rainfall increases slightly. Driving VECTRI with the model closest to the ensemble median keeps the projected malaria changes representative of the central climate change, but does not describe their spread; the rule-based approach, driven by all ten models, provides this spread for suitability. Both approaches agree that the changes are small by mid-century and under SSP2-4.5, and that transmission decreases under SSP5-8.5 at the end of the century, when temperatures exceed the thermal optimum for transmission [22] during more months and rainfall decreases by about one third. They also agree on a smaller decrease, or an increase, in the south-east, where the burden is concentrated.

For SMC, the main projection result is the stability of the seasonal timing: the centre of gravity of simulated transmission moves by less than 0.1 month and the share of the annual EIR in July-October remains about 90 % in all scenarios. The present calendar would therefore remain aligned with the climatic window of transmission, and the regional differences in observed timing, of more than one month between Kédougou and Casamance or the centre-west, matter more for planning than the projected changes. A regionally differentiated SMC calendar, starting earlier in the south-east and extending to November in the regions with late seasons, would be consistent with these results and with the protection decay observed after each round in Kédougou [46].

### Strengths and limitations

The strengths of this study are the use of two independent surveillance sources with near-universal testing, the harmonisation of district reports across formats, the evaluation of the climate forcing against observations, the use of ten bias-adjusted models and of an objective criterion to select the model driving VECTRI, the decomposition of the delay between rainfall and cases, and the use of resampling statistics without pairing simulated and observed years. Several limitations must be considered. CMIP6 historical simulations do not reproduce the observed chronology of years, so the comparison is restricted to climatological seasonality and spatial patterns and cannot assess interannual skill; driving VECTRI with observed or reanalysis forcing over the surveillance period would allow this evaluation. VECTRI was driven by a single model, and its parameters were not calibrated; the absolute EIR, the parasite ratio and the simulated cases are not realistic for current conditions, which include interventions, and the projections assume that interventions remain absent. The 0.25° grid represents Dakar by few cells, and the population is interpolated from 1°. Bias-adjusted rainfall is 10-30 % wetter than CHIRPS in the centre and west, which may lengthen the simulated season there. The two observed sources differ in 2022, although the main result did not depend on this year, and 2025 data are provisional. The comparison of departments relies on the assignment of districts to departments, whose boundaries differ. Finally, SMC coverage, indoor residual spraying and case management were not available by month and district and were not analysed.

## Conclusions

Confirmed malaria in Senegal follows a regular seasonal cycle centred on mid-October, with a burden increasingly concentrated in the south-east. VECTRI driven by bias-adjusted CMIP6 climate reproduces the timing of transmission and its spatial gradient: reported cases lag simulated transmission by about one month, and simulated transmission lags rainfall by about one month. The model does not reproduce the regional differences in timing or the distribution of clinical cases, which depend on hydrology, immunity, population and the health system. The TPR from district reports is a robust indicator for the evaluation of such models. The projected timing of transmission is stable under all scenarios, while its intensity decreases strongly only under SSP5-8.5 at the end of the century. Driving VECTRI with observed forcing, calibrating it against the TPR, including interventions and verifying the 2025 data are the next steps towards climate-informed planning of SMC and early warning.

## Data Availability

NEX-GDDP-CMIP6 and CHIRPS are publicly available. The aggregated results by region and department and the code of the analysis chain are available in the online dashboard (https://huggingface.co/spaces/idiouf/CLIMATE_MALARIA_SENEGAL_VECTRI_MODELING). District surveillance data belong to the National Malaria Control Programme (PNLP) of Senegal and are available on request to the PNLP.

## Acknowledgments

The authors thank the National Malaria Control Programme (PNLP) of Senegal for the surveillance data, the National Agency of Statistics and Demography (ANSD) for the population data, NASA for NEX-GDDP-CMIP6, the Climate Hazards Center for CHIRPS, the climate modelling groups of CMIP6 and the developers of VECTRI.

## Data availability

NEX-GDDP-CMIP6 and CHIRPS are publicly available. The aggregated results by region and department and the code of the analysis chain are available in the online dashboard [41]. District surveillance data belong to the PNLP and are available on request to the PNLP.

## Funding

This work was supported by Wellcome through the ACCLIMATISE project (Attribution of a Changing Climate in the Assessment of Malaria Intervention Strategy Efficiency; grant 308964/Z/23/Z). The funders had no role in study design, data collection and analysis, decision to publish, or preparation of the manuscript.

## Competing interests

The authors have declared that no competing interests exist.

## Author contributions

Conceptualization: Ibrahima Diouf. Data curation: Ibrahima Diouf, Elhadji Doucouré. Formal analysis: Ibrahima Diouf. Methodology: Ibrahima Diouf, Papa Fall. Resources: Elhadji Doucouré. Software: Ibrahima Diouf. Supervision: Amadou Thierno Gaye. Validation: Marie Jeanne Sambou Cissokho, Papa Fall, Amanatou Bèye Gueye. Visualization: Ibrahima Diouf. Writing - original draft: Ibrahima Diouf. Writing - review & editing: Ibrahima Diouf, Marie Jeanne Sambou Cissokho, Papa Fall, Amanatou Bèye Gueye, Elhadji Doucouré, Amadou Thierno Gaye.

## Supporting information

**S1 Fig. Annual confirmed cases in the two PNLP sources, 2016-2025:** regional database and harmonised district reports.

**S2 Fig. Lorenz curve of confirmed malaria cases against population across the 14 regions, 2009-2024.** The Gini coefficient is twice the area between the curve and the diagonal.

**S3 Fig. Annual incidence of confirmed malaria by region, 2009-2024**, with Sen slope (dashed) and Mann-Kendall statistics; 2025 (provisional, orange) not included in the trend.

**S4 Fig. Seasonal centre of gravity of confirmed cases by region and year, 2009-2024.** Circular mean month (9 = mid-September, 10 = mid-October, 11 = mid-November).

**S5 Fig. Monthly EIR simulated by VECTRI driven by MRI-ESM2-0, July to December, 1995-2014** (log classes).

**S6 Fig. Seasonal cycles by region.** Monthly share of the annual total of confirmed cases (PNLP, 2009-2024), EIR and new cases simulated by VECTRI (1995-2014) and CHIRPS rainfall (bars, 2009-2024). Mean incidence 2009-2024 in brackets.

**S7 Fig. Cross-correlation between the normalised seasonal cycles of confirmed cases (2009-2024) and simulated EIR (1995-2014) by region**, for lags of 0 to 4 months of cases behind the EIR; bold: maximum of each row.

**S8 Fig. Projected changes by region and model under SSP5-8.5, 2081-2100:** (a) annual rainfall (%) and (b) mean temperature (°C), relative to 1995-2014.

**S9 Fig. Rule-based climatic suitability, ten-model median, 1995-2014:** (a) suitable season length and (b) number of months with mean temperature above 34 °C.

## Notes

### Competing Interest Statement

The authors have declared no competing interest.

### Author Declarations

This study used only aggregated, anonymised routine surveillance data (monthly counts by health district and region) provided by the National Malaria Control Programme (PNLP) of Senegal. No individual patient data were collected or analysed. No IRB approval was required as the study did not involve direct human subjects research. Data were provided by the PNLP under a data sharing agreement for research purposes.

